# Reply to Dages, et al. You AIn’t using it right: Artificial intelligence progress in allergy

**DOI:** 10.1101/2023.08.14.23293937

**Authors:** Dylan Lawless

## Abstract

We present our playful yet serious response to Dages et al.’s letter on AI limitations in allergy, specifically the AI model, ChatGPT. Our lighthearted title, “*You AIn’t using it right: Artificial intelligence progress in allergy*,” belies our intent to show how AI tools can be used differently to yield reliable, factual information that can enhance the field of medical sciences. We demonstrate by using AI to build a computer program to download and analyse PubMed sources, summarise the content, provide supporting primary evidence, and produce figures to enlighten the reader. We believe that the correct and increasingly automated application of AI can revolutionize its use in medicine, catalyzing its evolution to deliver significant benefits. Therefore, while caution is required, we should not default to dismissing the power of these new tools.

## Main

We (ChatGPT and I) read with interest the letter by Dages et al., regarding the limitations of the AI tool ChatGPT [1]. We concur that the tool did not accurately address their query on cefazolin administration in a penicillin-allergic patient. However, we believe this outcome reflects the current limitations of the tool’s use as a search engine rather than the AI model itself. We fully agree that their submission is typical and acknowledge that results can often deviate convincingly from what is expected. Their report is thus an important cautionary example.

Since language models generate responses based on data likelihood, not factual accuracy, it is important that we understand their function to use them effectively.

To demonstrate, we used ChatGPT v4 for a more complex approach to answer the author’s same question. We propose that one must (for now) explicitly ask queries such as “Can we automate downloads from PubMed, analyze the content, and interpret the result concerning the safety of cefazolin for penicillin-allergic patients?”.

We began by asking “what R package is able to query PubMed and return abstracts?” (R being our programming language of choice). ChatGPT automatically generated R code which could gather relevant abstracts from PubMed using our keywords. From here we incrementally asked for additional code to process and interpret the information.

The full code set provided us with an R computer program, written by ChatGPT, which performs the automated PubMed queries relative to the question and saves all abstracts and references (supplemental). It then performs term-frequency and correlation analysis on the texts. Lastly, the text corpus is used as input for ChatGPT’s interpretation of the original question to return a response based on the referenced summaries (supplemental). The result intuitively demonstrates, to the human reader, what the text-based information consisted of before being interpreted by ChatGPT’s model.

The final result consists of **Figure 1 (A-D)** illustrating the summarized content from PubMed, **(E)** a balanced text report based on those summaries, and **(F)** the list of publications used in the analysis.

**Figure 1.**
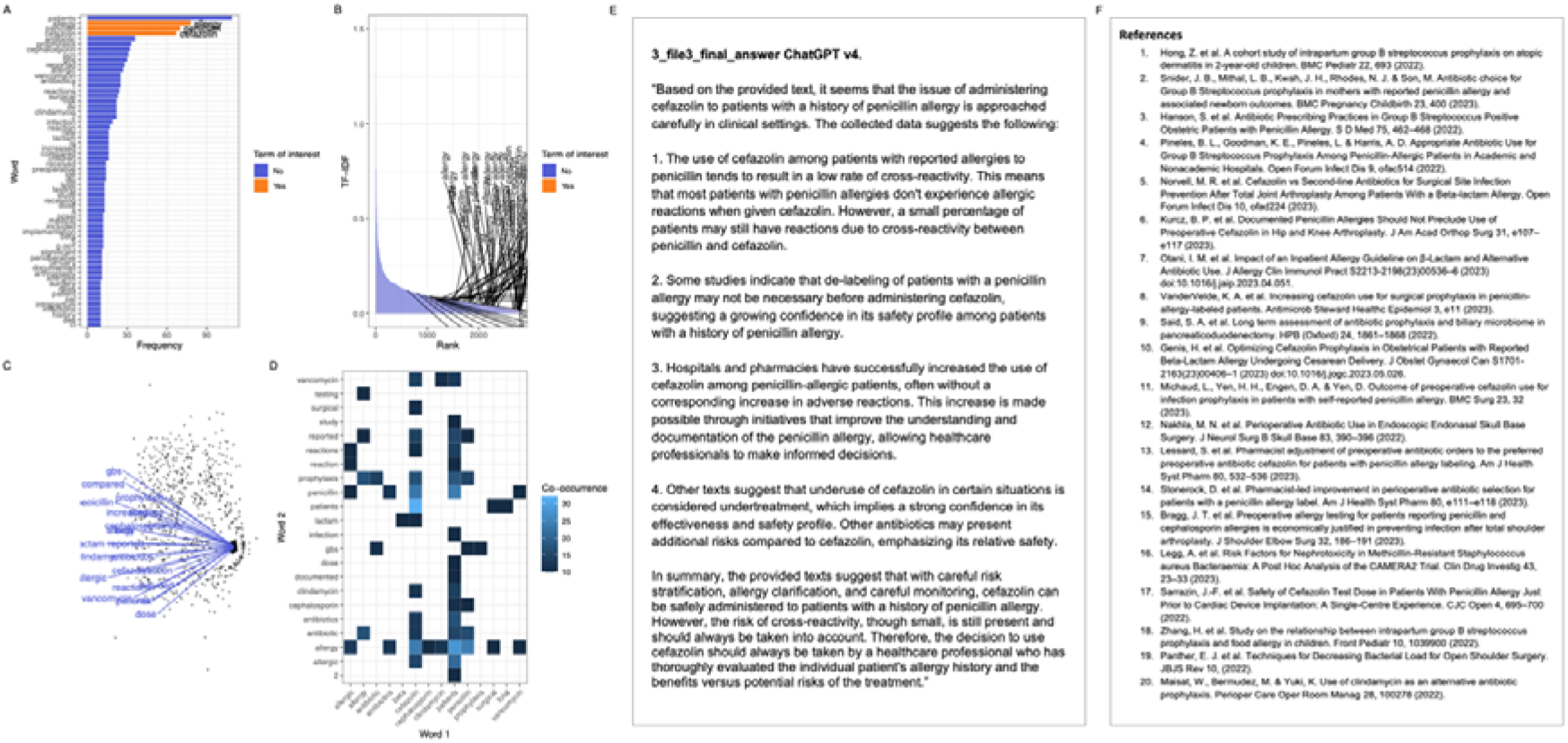
Term frequency in automated PubMed queries and final interpretation. **[A]** Most common words in abstracts related to cefazolin and penicillin allergy (quantile 0.95 shown). **[B]** TF-IDF for terms of interest in abstracts related to “cefazolin” and “penicillin” “allergy” (text labels highlight matches to the three terms). **[C]** Network plot of edge weights based on the frequency of co-occurrence in abstracts related to cefazolin and penicillin allergy (quantile 0.98 shown). **[D]** Heatmap of term co-occurrence in abstracts (co-occurrence threshold >9 shown). This same dataset throughout was used for subsequent interpretation by ChatGPT. **[E]** Final response by ChatGPT based on summaries from abstracts. **[F]** References used for ChatGPT’s final response. TF-IDF, term frequency-inverse document frequency.

The supporting data was automatically sourced for the first twenty PubMed results, the URLs and PMIDs printed and automatically imported into a citation manager. We limited the method to the default of twenty publication abstracts so that it is easy to replicate, however using ChatGPT API the system could be automated, with relative ease, to run on far larger datasets.

Our findings show that AI can provide valuable information for problem solving. We believe our colleagues’ concerns are justified since most users initially assume that responses to queries are supposed to be fact-based. By adjusting the approach, we can use AI tools to improve how we find and analyse information.

However, we must highlight that the process, as it stands, is time-intensive - our session required two hours. It would be unrealistic to expect every user to dedicate such effort. We anticipate the advent of more user-friendly tools and interfaces, making data retrieval and analysis accessible and efficient.

## Data Availability

All data produced in the present work are contained in the manuscript

