## Supplemental for "Reply to Dages, et al. You AIn’t using it right: Artificial intelligence progress in allergy": figure_1.pdf

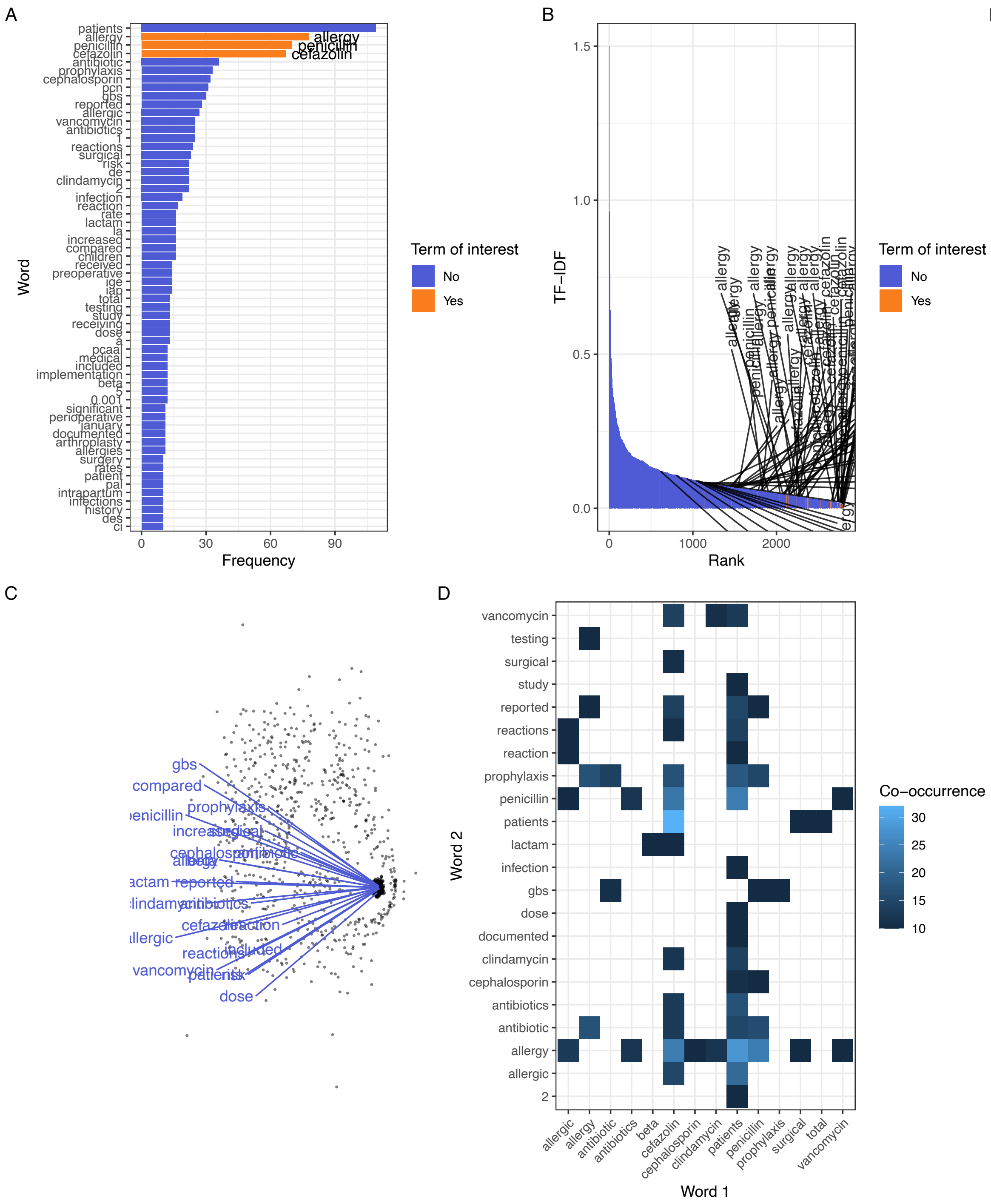


E

**3_file3_final_answer ChatGPT v4.**

“Based on the provided text, it seems that the issue of administering cefazolin to patients with a history of penicillin allergy is approached carefully in clinical settings. The collected data suggests the following:

1. The use of cefazolin among patients with reported allergies to penicillin tends to result in a low rate of cross-reactivity. This means that most patients with penicillin allergies don't experience allergic reactions when given cefazolin. However, a small percentage of patients may still have reactions due to cross-reactivity between penicillin and cefazolin.

2. Some studies indicate that de-labeling of patients with a penicillin allergy may not be necessary before administering cefazolin, suggesting a growing confidence in its safety profile among patients with a history of penicillin allergy.

3. Hospitals and pharmacies have successfully increased the use of cefazolin among penicillin-allergic patients, often without a corresponding increase in adverse reactions. This increase is made possible through initiatives that improve the understanding and documentation of the penicillin allergy, allowing healthcare professionals to make informed decisions.

4. Other texts suggest that underuse of cefazolin in certain situations is considered undertreatment, which implies a strong confidence in its effectiveness and safety profile. Other antibiotics may present additional risks compared to cefazolin, emphasizing its relative safety.

In summary, the provided texts suggest that with careful risk stratification, allergy clarification, and careful monitoring, cefazolin can be safely administered to patients with a history of penicillin allergy. However, the risk of cross-reactivity, though small, is still present and should always be taken into account. Therefore, the decision to use cefazolin should always be taken by a healthcare professional who has thoroughly evaluated the individual patient's allergy history and the benefits versus potential risks of the treatment.”

F
