## Supplemental for "Reply to Dages, et al. You AIn’t using it right: Artificial intelligence progress in allergy": 5_pubmed_references.docx

1. Hong, Z. et al. A cohort study of intrapartum group B streptococcus prophylaxis on atopic dermatitis in 2-year-old children. BMC Pediatr 22, 693 (2022).
2. Snider, J. B., Mithal, L. B., Kwah, J. H., Rhodes, N. J. & Son, M. Antibiotic choice for Group B Streptococcus prophylaxis in mothers with reported penicillin allergy and associated newborn outcomes. BMC Pregnancy Childbirth 23, 400 (2023).
3. Hanson, S. et al. Antibiotic Prescribing Practices in Group B Streptococcus Positive Obstetric Patients with Penicillin Allergy. S D Med 75, 462–468 (2022).
4. Pineles, B. L., Goodman, K. E., Pineles, L. & Harris, A. D. Appropriate Antibiotic Use for Group B Streptococcus Prophylaxis Among Penicillin-Allergic Patients in Academic and Nonacademic Hospitals. Open Forum Infect Dis 9, ofac514 (2022).
5. Norvell, M. R. et al. Cefazolin vs Second-line Antibiotics for Surgical Site Infection Prevention After Total Joint Arthroplasty Among Patients With a Beta-lactam Allergy. Open Forum Infect Dis 10, ofad224 (2023).
6. Kurcz, B. P. et al. Documented Penicillin Allergies Should Not Preclude Use of Preoperative Cefazolin in Hip and Knee Arthroplasty. J Am Acad Orthop Surg 31, e107–e117 (2023).
7. Otani, I. M. et al. Impact of an Inpatient Allergy Guideline on β-Lactam and Alternative Antibiotic Use. J Allergy Clin Immunol Pract S2213-2198(23)00536–6 (2023) doi:10.1016/j.jaip.2023.04.051.
8. VanderVelde, K. A. et al. Increasing cefazolin use for surgical prophylaxis in penicillin-allergy-labeled patients. Antimicrob Steward Healthc Epidemiol 3, e11 (2023).
9. Said, S. A. et al. Long term assessment of antibiotic prophylaxis and biliary microbiome in pancreaticoduodenectomy. HPB (Oxford) 24, 1861–1868 (2022).
10. Genis, H. et al. Optimizing Cefazolin Prophylaxis in Obstetrical Patients with Reported Beta-Lactam Allergy Undergoing Cesarean Delivery. J Obstet Gynaecol Can S1701-2163(23)00406–1 (2023) doi:10.1016/j.jogc.2023.05.026.
11. Michaud, L., Yen, H. H., Engen, D. A. & Yen, D. Outcome of preoperative cefazolin use for infection prophylaxis in patients with self-reported penicillin allergy. BMC Surg 23, 32 (2023).
12. Nakhla, M. N. et al. Perioperative Antibiotic Use in Endoscopic Endonasal Skull Base Surgery. J Neurol Surg B Skull Base 83, 390–396 (2022).
13. Lessard, S. et al. Pharmacist adjustment of preoperative antibiotic orders to the preferred preoperative antibiotic cefazolin for patients with penicillin allergy labeling. Am J Health Syst Pharm 80, 532–536 (2023).
14. Stonerock, D. et al. Pharmacist-led improvement in perioperative antibiotic selection for patients with a penicillin allergy label. Am J Health Syst Pharm 80, e111–e118 (2023).
15. Bragg, J. T. et al. Preoperative allergy testing for patients reporting penicillin and cephalosporin allergies is economically justified in preventing infection after total shoulder arthroplasty. J Shoulder Elbow Surg 32, 186–191 (2023).
16. Legg, A. et al. Risk Factors for Nephrotoxicity in Methicillin-Resistant Staphylococcus aureus Bacteraemia: A Post Hoc Analysis of the CAMERA2 Trial. Clin Drug Investig 43, 23–33 (2023).
17. Sarrazin, J.-F. et al. Safety of Cefazolin Test Dose in Patients With Penicillin Allergy Just Prior to Cardiac Device Implantation: A Single-Centre Experience. CJC Open 4, 695–700 (2022).
18. Zhang, H. et al. Study on the relationship between intrapartum group B streptococcus prophylaxis and food allergy in children. Front Pediatr 10, 1039900 (2022).
19. Panther, E. J. et al. Techniques for Decreasing Bacterial Load for Open Shoulder Surgery. JBJS Rev 10, (2022).
20. Maisat, W., Bermudez, M. & Yuki, K. Use of clindamycin as an alternative antibiotic prophylaxis. Perioper Care Oper Room Manag 28, 100278 (2022).
