## Supplemental for "Reply to Dages, et al. You AIn’t using it right: Artificial intelligence progress in allergy": 6_jaci_gpt_chat.docx

This document contains the conversation with ChatGPT v4 for “*You AIn’t using it right: Artificial intelligence progress in allergy”.*

**User**

what R package is able to query pubmed and return abstracts

**ChatGPT**

There are a few R packages that can be used to query PubMed and return abstracts. Here are a couple:

1. ```rentrez```: This package is an R interface to the NCBI's EUtils API, which provides access to PubMed and other databases. Here's an example of how you can use this package to get abstracts:

```r

library(rentrez)

\# Search for publications about "CRISPR" in PubMed

papers <- entrez_search(db="pubmed", term="CRISPR")

\# Fetch records in "abstract" format

abstracts <- entrez_fetch(db="pubmed", id=papers$ids, rettype="abstract")

```

2. ```easyPubMed```: This package simplifies the process of downloading PubMed records and converting them into R-friendly formats. Here's an example of how to use it:

```r

library(easyPubMed)

\# Define a query

query <- '("CRISPR"[TI]) AND ("2023"[DP])'

\# Fetch records

records <- get_pubmed_ids(query)

\# Fetch abstracts

abstracts <- fetch_pubmed_data(records, format = 'abstract')

```

Please note that these examples are simplified. In a real scenario, you would need to deal with things like error checking, pagination for large result sets, and possibly rate limiting. Be sure to refer to the documentation of the packages and the NCBI EUtils API for more details.

**User**

Great. We can work with the first package, rentrez. Next, on pubmed, here an example of the structure of a URL after I query three terms. Can you identify the three terms such that we could make a variable to search for similar combinations ? ```https://www.ncbi.nlm.nih.gov/pmc?term=((cefazolin%5BAbstract%5D)%20AND%20penicillin%5BAbstract%5D)%20AND%20allergy%5BAbstract%5D```

**ChatGPT**

Sure. In the URL you have provided, the terms are 'cefazolin', 'penicillin', and 'allergy'. The '%5BAbstract%5D' is URL encoding for '[Abstract]', specifying that the search should be limited to the abstract of the articles. 'AND' is used to combine the search terms.

Here's how you could create a similar query using the ```rentrez``` package:

```r

library(rentrez)

\# Define your search terms

terms <- c("cefazolin", "penicillin", "allergy")

\# Create the query string by joining the terms with '[Abstract] AND '

query <- paste0(terms, "[Abstract]", collapse = " AND ")

\# Search for publications in PubMed

papers <- entrez_search(db="pubmed", term=query)

\# Fetch records in "abstract" format

abstracts <- entrez_fetch(db="pubmed", id=papers$ids, rettype="abstract")

```

This script will search for articles in which the abstracts contain all of the terms 'cefazolin', 'penicillin', and 'allergy'.

**User**

Great, however the text block that is created after this query is very big. Can we split it into smaller chunks such that it is easy to define where each abstract starts and ends and then save them to file?

**ChatGPT**

Yes, you're right. The ```entrez_fetch``` function returns all the abstracts concatenated into one big character string. We can make this more manageable by using the ```entrez_summary``` function instead, which returns a list of summary records.

Here's how you can modify the script to store each abstract separately:

```r

library(rentrez)

\# Define your search terms

terms <- c("cefazolin", "penicillin", "allergy")

\# Create the query string by joining the terms with '[Abstract] AND '

query <- paste0(terms, "[Abstract]", collapse = " AND ")

\# Search for publications in PubMed

papers <- entrez_search(db="pubmed", term=query)

\# Fetch summaries

summaries <- entrez_summary(db="pubmed", id=papers$ids)

\# Extract the abstracts

abstracts <- sapply(summaries, function(summary) summary$abstract)

\# Write each abstract to a separate file

for (i in seq_along(abstracts)) {

file_name <- paste0("abstract_", i, ".txt")

writeLines(abstracts[i], file_name)

}

```

This script writes each abstract to a separate text file named "abstract_1.txt", "abstract_2.txt", etc.

Please note that the ```entrez_summary``` function only returns a summary of each article, and the amount of information included in the summary can vary. If the abstract is not included in the summary, you will need to fetch the full record instead. However, this involves more complex parsing of the returned XML or ASN.1 data.

Additionally, be aware that there are usage policies and guidelines for E-utilities API which includes an upper limit to the number of requests per second and a recommendation to avoid excessive server load (https://www.ncbi.nlm.nih.gov/books/NBK25497/#_chapter2_Usage_Guidelines_and_Requirem_). Be sure to respect these in your scripts.

**User**

this is good, however ```> # Write each abstract to a separate file

> for (i in seq_along(abstracts)) {

+	file_name <- paste0("abstract_", i, ".txt")

+	writeLines(abstracts[i], file_name)

+ }

Error in writeLines(abstracts[i], file_name) :

can only write character objects``` - I have tried ```for (i in seq_along(abstracts)) {

file_name <- paste0("../data/abstract_", i, ".txt")

writeLines(as.character(abstracts[i]), file_name)

}

`` but this prints "NULL" to each file.

**ChatGPT**

I'm sorry for the confusion. It seems I made a mistake in my response. The function ```entrez_summary``` does not return the abstract of the articles. We have to stick to ```entrez_fetch``` and parse the result.

The returned object from ```entrez_fetch``` with ```rettype="abstract"``` is a large text block with XML structure. We need to parse this XML to extract the abstracts. The ```XML``` package in R can help with this. Here's how you can modify the script:

```r

library(rentrez)

library(XML)

\# Define your search terms

terms <- c("cefazolin", "penicillin", "allergy")

\# Create the query string by joining the terms with '[Abstract] AND '

query <- paste0(terms, "[Abstract]", collapse = " AND ")

\# Search for publications in PubMed

papers <- entrez_search(db="pubmed", term=query)

\# Fetch records in "abstract" format

xmlData <- entrez_fetch(db="pubmed", id=papers$ids, rettype="abstract", retmode="xml")

\# Parse XML

doc <- xmlParse(xmlData)

\# Extract abstract nodes

abstractNodes <- getNodeSet(doc, "//AbstractText")

\# Extract the content of abstract nodes

abstracts <- sapply(abstractNodes, xmlValue)

\# Write each abstract to a separate file

for (i in seq_along(abstracts)) {

file_name <- paste0("abstract_", i, ".txt")

writeLines(abstracts[i], file_name)

}

```

This script parses the XML returned by ```entrez_fetch```, extracts the abstracts, and writes each one to a separate file. Make sure you have the XML package installed in R (you can install it with ```install.packages("XML")``` if you haven't).

**User**

this is great. can we instead print it to a single file

**ChatGPT**

Yes, you can certainly write all abstracts to a single file. A simple way to do this could be to concatenate all the abstracts into a single character string, with each abstract separated by a couple of newline characters for readability. Here's how you can do it:

```r

library(rentrez)

library(XML)

\# Define your search terms

terms <- c("cefazolin", "penicillin", "allergy")

\# Create the query string by joining the terms with '[Abstract] AND '

query <- paste0(terms, "[Abstract]", collapse = " AND ")

\# Search for publications in PubMed

papers <- entrez_search(db="pubmed", term=query)

\# Fetch records in "abstract" format

xmlData <- entrez_fetch(db="pubmed", id=papers$ids, rettype="abstract", retmode="xml")

\# Parse XML

doc <- xmlParse(xmlData)

\# Extract abstract nodes

abstractNodes <- getNodeSet(doc, "//AbstractText")

\# Extract the content of abstract nodes

abstracts <- sapply(abstractNodes, xmlValue)

\# Combine all abstracts into a single string, separated by two newline characters

all_abstracts <- paste(abstracts, collapse = "\n\n")

\# Write all abstracts to a single file

writeLines(all_abstracts, "all_abstracts.txt")

```

This script will create a file named "all_abstracts.txt", where each abstract is separated by a blank line.

**User**

Great, now with these abstracts, we are going to read the text and based on the content we will ask "is it safe to give cefazolin to a patient with a history of penicillin allergy?” - however we do not want external information that you may know - we only want to see what facts are present in the following text. We will also iterate through several chucks:

```

Cefazolin is a first-line agent for prevention of surgical site infections (SSIs) after total joint arthroplasty. Patients labeled allergic to beta-lactam antibiotics frequently receive clindamycin or vancomycin perioperatively due to the perceived risk of a hypersensitivity reaction after exposure to cefazolin.

This single-system retrospective review included patients labeled allergic to penicillin or cephalosporin antibiotics who underwent a primary total hip and/or knee arthroplasty between January 2020 and July 2021. A detailed chart review was performed to compare the frequency of SSI within 90 days of surgery and interoperative hypersensitivity reactions (HSRs) between patients receiving cefazolin and patients receiving clindamycin and/or vancomycin.

A total of 1128 hip and/or knee arthroplasties from 1047 patients were included in the analysis (cefazolin n = 809, clindamycin/vancomycin n = 319). More patients in the clindamycin and/or vancomycin group had a history of cephalosporin allergy and allergic reactions with immediate symptoms. There were fewer SSIs in the cefazolin group compared with the clindamycin and/or vancomycin group (0.9% vs 3.8%; P < .001) including fewer prosthetic joint infections (0.1% vs 1.9%). The frequency of interoperative HSRs was not different between groups (cefazolin = 0.2% vs clindamycin/vancomycin = 1.3%; P = .06).

The use of cefazolin as a perioperative antibiotic for infection prophylaxis in total joint arthroplasty in patients labeled beta-lactam allergic is associated with decreased postoperative SSI without an increase in interoperative HSR.

To evaluate the choice of antibiotic used for intrapartum Group B Streptococcus (GBS) prophylaxis in pregnant individuals with reported penicillin allergies compared to those without reported penicillin allergies and investigate whether there are associated differences in neonatal outcomes.

This retrospective cohort study included mother-infant dyads of GBS positive pregnant individuals who labored and delivered newborns ≥ 35 weeks of gestation at a high-volume urban hospital (2005-2018). The type of antibiotic administered to the mothers for GBS prophylaxis (beta-lactam prophylaxis defined as penicillin-class drug or cefazolin; alternative prophylaxis defined as vancomycin or clindamycin) was compared between those with a penicillin allergy documented in their medical record versus those who did not. Neonatal outcomes included number of postnatal blood draws, antibiotic administration, neonatal intensive care unit (NICU) admission, bacteremia, and hospital length of stay and were compared between groups. Bivariable and multivariable analyses were performed.

Of 11,334 mother-infant pairs, 1170 (10.3%) mothers had a penicillin allergy documented in their medical record. Of them, 49 (4.2%) received a penicillin, 259 (22.1%) received cefazolin, 449 (38.4%) received clindamycin, and 413 (35.3%) received vancomycin. Patients with a reported penicillin allergy were significantly more likely to receive alternative GBS prophylaxis compared to those without penicillin allergy (73.7% vs. 0.2%, p < 0.01). Neonates of patients who received alternative GBS prophylaxis were significantly more likely to undergo a postnatal lab draw compared to neonates of patients who received beta-lactam antibiotics (20.8% vs. 17.3%, OR 1.25 (95% CI 1.08-1.46)). This significant association persisted after adjusting for potential confounders (aOR 1.23, 95% CI 1.06-1.43). There were no other significant differences seen in other newborn outcomes.

```

**ChatGPT**

Based on the given abstracts, here are the relevant points that can help answer the question "Is it safe to give cefazolin to a patient with a history of penicillin allergy?"

1. In a retrospective review involving patients labeled allergic to penicillin or cephalosporin antibiotics who underwent a primary total hip and/or knee arthroplasty, there were fewer surgical site infections (SSIs) in the cefazolin group compared with the clindamycin and/or vancomycin group. Importantly, the frequency of intraoperative hypersensitivity reactions (HSRs) was not significantly different between the groups (cefazolin = 0.2% vs clindamycin/vancomycin = 1.3%; P = .06).

Conclusion: The use of cefazolin as a perioperative antibiotic in these surgical procedures in patients labeled as beta-lactam allergic was associated with a decreased rate of postoperative SSIs without an increase in intraoperative HSR.

2. In a retrospective cohort study of Group B Streptococcus (GBS) prophylaxis in pregnant individuals, 259 out of 1170 (22.1%) mothers with a documented penicillin allergy received cefazolin. The study showed that neonates of patients who received alternative GBS prophylaxis (vancomycin or clindamycin) were significantly more likely to undergo a postnatal lab draw compared to neonates of patients who received beta-lactam antibiotics (including cefazolin), but there were no other significant differences in newborn outcomes.

Conclusion: Cefazolin was administered to a significant proportion of mothers with a documented penicillin allergy for GBS prophylaxis during pregnancy, and there were no mentioned adverse outcomes specific to this group.

Based on these abstracts, cefazolin was administered to patients with a history of penicillin allergy in different scenarios and did not demonstrate an increase in hypersensitivity reactions or other adverse outcomes compared to alternative antibiotics. However, these are abstracts and the full studies might contain more nuanced information or limitations.

**User**

Great, now with these abstracts, we are going to read the text and based on the content we will ask "is it safe to give cefazolin to a patient with a history of penicillin allergy?” - however we do not want external information that you may know - we only want to see what facts are present in the following text. We will also iterate through several chucks:

```

Pregnant individuals who report a penicillin allergy were more likely to receive alternative antibiotics for GBS prophylaxis compared to those without a penicillin allergy. This was associated with an increased frequency of postnatal blood draws among neonates of mothers with a reported penicillin allergy. Administration of alternative intrapartum antibiotic prophylaxis with vancomycin or clindamycin is common in individuals with self-reported penicillin allergy, and maternal alternative antibiotic administration may impact neonatal care, particularly via increased lab draws.

Evaluate the impact of an allergy history-guided algorithm for optimizing perioperative cefazolin use in patients with reported beta-lactam allergy undergoing cesarean delivery.

The Allergy Clarification for Cefazolin Evidence-based Prescribing Tool (ACCEPT) was developed through consensus by allergists, anesthesiologists, and infectious diseases specialists, and implemented over a 2-month period (December 1, 2018, to January 31, 2019). A segmented regression on monthly cefazolin use was conducted during the baseline (January 1 to November 30, 2018) and intervention (February 1 to December 31, 2019) periods to evaluate the impact of ACCEPT on the monthly use of perioperative cefazolin in patients with reported beta-lactam allergy undergoing cesarean delivery. The frequency of perioperative allergic reactions and surgical site infections was collected during both periods.

Of the 3128 eligible women who underwent a cesarean delivery, 282 (9%) reported a beta-lactam allergy. The most common beta-lactam allergens were penicillin (64.3%), amoxicillin (16.0%), and cefaclor (6.0%). The most frequently reported allergic reactions were rash (38.1%), hives (21.4%), and unknown (11.6%). Use of cefazolin increased from 52% (baseline) to 87% during the intervention period. Segmented regression analysis confirmed a statistically significant increase following implementation (incidence rate ratio 1.62, 95% CI 1.19 - 2.21, P = 0.002). There was 1 perioperative allergic reaction in the baseline period and 2 during the intervention period. Cefazolin use remained high (92%) 2 years after algorithm implementation.

Implementation of a simple allergy history-guided algorithm in obstetrical patients with reported beta-lactam allergy resulted in a sustained increase in perioperative cefazolin prophylaxis.

A guideline identifying when inpatients with penicillin or cephalosporin antibiotic allergy labels (PCAAL) can receive β-lactam antibiotics increased β-lactam receipt at a large northeastern US health care system.

To report outcomes of implementing a similar guideline and electronic order set (OS) at an independent academic health care system.

Penicillin/cephalosporin receipt (percentage of inpatients receiving full doses) and alternative antibiotic use (days of therapy per 1000 patient-days [DOT/1000PD]) were compared over 3 periods before (February 1, 2017, to January 31, 2018) and after guideline implementation (February 1, 2018, to January 31, 2019), and after OS implementation (February 1, 2019, to January 31, 2020) among inpatients with PCAAL admitted on medical services with access to guideline/OS and education (Medical-PCAAL, n = 8721), surgical services with access to guideline/OS without education (Surgical-PCAAL, n = 5069), and obstetrics/gynecology services without interventions (Ob/Gyn-PCAAL, n = 798) and inpatients without PCAAL admitted on the same services (Medical-No-PCAAL, n = 50,840; Surgical-No-PCAAL, n = 29,845; Ob/Gyn-No-PCAAL, n = 6109). χ2 tests were used to compare categorical variables, and analysis of variance was used to compare continuous and interrupted time series analyses (ITSA) to investigate the guideline/OS implementation effect on penicillin/cephalosporin receipt.

In the Medical-PCAAL group, penicillin/cephalosporin receipt increased (58%-68%, P < .001), specifically for cefazolin (8%-11%, P = .02) and third- to fifth-generation cephalosporins (43%-48%, P = .04), and aztreonam use decreased (12 DOT/1000PD, P = .03). In the Medical-No-PCAAL group, penicillin/cephalosporin receipt increased (88%-90%, P = .004), specifically for penicillin (40%-44%, P < .001), without changes in aztreonam use. Significant changes were not observed in these outcomes on surgical or obstetrics/gynecology services. Per ITSA, guideline/OS implementation was associated with increased penicillin/cephalosporin receipt in the Medical-PCAAL group only.

```

**ChatGPT**

Based on the given abstracts, here are the relevant points that can help answer the question "Is it safe to give cefazolin to a patient with a history of penicillin allergy?"

1. A study of pregnant individuals who reported a penicillin allergy revealed that they were more likely to receive alternative antibiotics for GBS prophylaxis, which led to an increased frequency of postnatal blood draws for their neonates. However, the abstract does not specifically discuss outcomes related to the administration of cefazolin in these cases.

2. The Allergy Clarification for Cefazolin Evidence-based Prescribing Tool (ACCEPT) was developed and implemented to guide the use of perioperative cefazolin in patients with reported beta-lactam allergies undergoing cesarean delivery. Of the eligible women, 9% reported a beta-lactam allergy, with penicillin being the most common allergen. The use of cefazolin increased significantly from 52% (baseline) to 87% during the intervention period. Notably, there was only a small increase in perioperative allergic reactions (1 in the baseline period and 2 during the intervention period), suggesting that the administration of cefazolin was generally safe.

3. The implementation of a guideline identifying when inpatients with penicillin or cephalosporin antibiotic allergy labels (PCAAL) can receive β-lactam antibiotics led to an increase in β-lactam receipt. In the Medical-PCAAL group, the use of cefazolin specifically increased (from 8% to 11%), and there were no significant changes in these outcomes on surgical or obstetrics/gynecology services. The abstract does not mention any increase in allergic reactions or adverse outcomes associated with this increase in cefazolin use.

Based on these abstracts, it appears that in certain clinical situations, cefazolin was safely administered to patients with a history of penicillin allergy without a significant increase in allergic reactions or adverse outcomes. However, these are abstracts and the full studies might contain more nuanced information or limitations.

**User**

Great, now with these abstracts, we are going to read the text and based on the content we will ask "is it safe to give cefazolin to a patient with a history of penicillin allergy?” - however we do not want external information that you may know - we only want to see what facts are present in the following text. We will also iterate through several chucks:

```

Guideline and OS implementation was associated with improved antibiotic stewardship on inpatient services that also received allergy education.

Group B Streptococcus (GBS) positive patients with penicillin allergies receive intrapartum antibiotics for neonatal sepsis prophylaxis based on recommendations from the American College of Obstetricians and Gynecologists (ACOG). The objective of this study was to determine which antibiotics are used in GBS positive patients with documented penicillin allergies and evaluate for antibiotic stewardship improvements at a tertiary hospital in the Midwestern U.S.

Retrospective chart review identified GBS positive patients with and without penicillin allergies admitted to the labor and delivery floor. EMR-documented penicillin allergy severity, results of antibiotic susceptibility testing, and all antibiotics administered from admission to delivery were recorded. The study population was divided based on penicillin allergy status with antibiotic choice analyzed using Fisher's exact test.

406 GBS positive patients underwent labor between May 1, 2019, and April 30, 2020. Penicillin allergy was documented in 62 (15.3 percent) patients. Of these patients, cefazolin and vancomycin were prescribed most frequently for intrapartum neonatal sepsis prophylaxis. Antibiotic susceptibility testing was performed on the GBS isolate in 74.2 percent of the penicillin allergic patients. Between penicillin allergy and no penicillin allergy groups, the frequency of ampicillin, cefazolin, clindamycin, gentamicin, and vancomycin use showed statistical differences.

The study results suggest that antibiotic choice for neonatal sepsis prophylaxis in GBS positive patients with penicillin allergy at a tertiary Midwestern hospital follows current ACOG guidelines. Cefazolin was used most frequently in this population followed by vancomycin and clindamycin. Our results identify room for improvement regarding regular antibiotic susceptibility testing in GBS positive patients with penicillin allergy.

Benzoyl peroxide (BPO) 5% has been shown to reduce Cutibacterium acnes load on the skin. BPO 5% with miconazole nitrate (MN) 2% may be beneficial, whereas BPO 5% with clindamycin cream 1% to 1.2% does not seem to have additive effects when compared with BPO 5% alone. Chlorhexidine gluconate solutions reduce the total bacterial load on the skin, but do not seem to have a significant effect on C. acnes.

ChloraPrep seems to be the best surgical skin preparation to decrease overall positive skin cultures. Preincisional hydrogen peroxide 3% application has been shown to be a cost-effective practice to inhibit growth of C. acnes. Vancomycin powder before deltopectoral interval closure has antimicrobial effects against C. acnes and is a cost-effective practice. Finally, Bactisure surgical lavage is protective against the formation of biofilms.

IV cefazolin has been shown to be more effective for shoulder arthroplasty infection prophylaxis than antibiotic alternatives such as vancomycin. Thus, patients with a questionable history of penicillin allergy should undergo additional testing.

For shoulder surgery infection prophylaxis, we recommend the use of BPO 5% cream for 5 days preoperatively with chlorhexidine wipes the night before and the morning of surgery. IV cefazolin should be administered perioperatively, and patients with a questionable history of penicillin allergy should be tested. Surgeons should consider preincisional application of hydrogen peroxide 3% for 5 minutes, followed by standard ChloraPrep preparation. Normal saline should be used for preclosure lavage. Finally, application of vancomycin powder deep to the deltopectoral interval closure should be considered.

Cephalosporins are the preferred antibiotics for prophylaxis against surgical site infections. Most studies give a rate of combined IgE and non-IgE penicillin allergy yet it is recommended that cephalosporins be avoided in patients having the former but can be used in those with the latter. Some studies use penicillin allergy while others penicillin family allergy rates. The primary goal of this study was to determine the rates of IgE and non-IgE allergy as well as cross reactions to both penicillin and the penicillin family. Secondary goals were to determine the surgical services giving preoperative cefazolin and the types of self reported reactions that patients' had to penicillin prompting their allergy status.

All patients undergoing elective and emergency surgery at a University Health Sciences Centre were retrospectively studied. The hospital electronic medical record was used for data collection.

```

**ChatGPT**

Based on the given abstracts, here are the relevant points that can help answer the question "Is it safe to give cefazolin to a patient with a history of penicillin allergy?"

1. A retrospective study aimed at determining which antibiotics were used for GBS positive patients with documented penicillin allergies found that cefazolin was frequently prescribed for intrapartum neonatal sepsis prophylaxis. While the study did not provide specific safety outcomes for the patients who received cefazolin, it indicated that the antibiotic choice followed the current guidelines of the American College of Obstetricians and Gynecologists (ACOG).

2. In relation to shoulder surgery infection prophylaxis, it was mentioned that IV cefazolin has been shown to be more effective than antibiotic alternatives such as vancomycin. Further, the text recommends that patients with a questionable history of penicillin allergy should undergo additional testing, potentially suggesting that cefazolin may be safe to use in some patients with a history of penicillin allergy, following further allergenic testing.

3. Cephalosporins, such as cefazolin, are presented as the preferred antibiotics for prophylaxis against surgical site infections. However, it is recommended that cephalosporins be avoided in patients with a history of IgE penicillin allergy but can be used in those with non-IgE penicillin allergy.

In conclusion, based on the given abstracts, it appears that cefazolin is frequently used and potentially safe for patients with a history of penicillin allergy, especially in cases with non-IgE penicillin allergy or those with a questionable history of allergy who undergo additional testing. As always, these are abstracts and the full studies might contain more nuanced information or limitations.

**User**

Great, now with these abstracts, we are going to read the text and based on the content we will ask "is it safe to give cefazolin to a patient with a history of penicillin allergy?” - however we do not want external information that you may know - we only want to see what facts are present in the following text. We will also iterate through several chucks:

```

8.9% of our patients reported non-IgE reactions to penicillin with a cross reactivity rate of 0.9% with cefazolin. 4.0% of our patients reported IgE reactions to penicillin with a cross reactivity rate of 4.0% with cefazolin. 10.5% of our patients reported non-IgE reactions to the penicillin family with a cross reactivity rate of 0.8% with cefazolin. 4.3% of our patients reported IgE reactions to the penicillin family with a cross reactivity rate of 4.0% with cefazolin.

Our rate of combined IgE and non-IgE reactions for both penicillin and penicillin family allergy was within the range reported in the literature. Our rate of cross reactivity between cefazolin and combined IgE and non-IgE allergy both to penicillin and the penicillin family were lower than reported in the old literature but within the range of the newer literature. We found a lower rate of allergic reaction to a cephalosporin than reported in the literature. We documented a wide range of IgE and non-IgE reactions. We also demonstrated that cefazolin is frequently the preferred antibiotics for prophylaxis against surgical site infections by many surgical services and that de-labelling patients with penicillin allergy is unnecessary.

Penicillin (PCN) allergy labels affect antimicrobial selection for surgical prophylaxis. We aimed to increase the percentage of cefazolin usage in patients with PCN allergy labels undergoing orthopedic surgery from 50% to 80%.

Quality improvement initiative.

Children's Mercy Kansas City (CMKC), a freestanding children's hospital.

Children scheduled for an orthopedic surgery (excluding spinal surgery) at CMKC who had a PCN allergy label and received a perioperative antibiotic.

No standardized process existed to identify and clarify PCN-allergic-labeled patients preoperatively. We developed a process for patient identification combined with a pharmacist phone interview for PCN allergy clarification. In plan-do-study-act (PDSA) part 1, we implemented a computer-generated patient list. In PDSA part 2, we combined automated identification with a phone interview. In PDSA part 3, we enhanced the patient list, making it timely and concise. In PDSA part 4, we included a PCN allergy clarification electronic survey to caregivers via the electronic medical record.

Cefazolin use in PCN-allergic surgical patients increased from 50% to 74% following interventions. Patients who had their PCN allergy label clarified were 4 times more likely to receive cefazolin compared to those whose allergy labels were not clarified (OR, 4.21; 95% CI, 1.68-11.61; P = 0.003). Moreover, 90% of patients received cefazolin when their PCN allergy was clarified and cefazolin was recommended. When a PCN allergy label was not clarified, only 59% of patients received cefazolin.

Appropriate clarification and documentation of PCN allergy labels increases the use of cefazolin for surgical prophylaxis.

Surgical patients with a penicillin allergy label (PAL) are less likely to receive β-lactams for surgical site infection (SSI) prophylaxis and more likely to receive second-line antibiotics, which may increase the risk of SSI, drug toxicities, and associated costs. We assessed the impact of implementing a pharmacist-led quality improvement project to increase the use of cefazolin as a first-line agent in this population.

After implementation of a pilot project in December 2021, all patients with a PAL and orders for preoperative antibiotics were risk stratified into high- or low-risk categories by a pharmacist. For the low-risk group, cefazolin was recommended. For the high-risk group, cefazolin was avoided and a second-line agent was administered. Our analysis compared 422 preintervention patients (August 15 to November 15, 2021) to 492 postintervention patients (December 15 to March 15, 2022). During the postintervention period, β-lactam usage increased (from 12.6% to 37.8%, P < 0.001), while usage of vancomycin (45.5% vs 29.5%, P < 0.001) and other second-line antibiotics (87.4% vs 62.2%, P < 0.001) declined. There were no adverse reactions reported in the preintervention cohort, with 2 potential adverse reactions reported after the intervention (0% vs 0.4%, P = 0.190). Medication costs based on claims data were 50% to 80% lower for patients receiving cefazolin.

```

**ChatGPT**

Based on the text provided, the following facts can be established regarding the safety of giving cefazolin to a patient with a history of penicillin allergy:

1. The cross-reactivity rate between cefazolin and both non-IgE and IgE reactions to penicillin was relatively low, at 0.9% and 4.0% respectively. Similar rates were observed for those who reported reactions to the penicillin family. The rates of cross reactivity were found to be lower than previously reported in older literature but consistent with more recent literature.

2. A report indicates that cefazolin is frequently the preferred antibiotic for prophylaxis against surgical site infections, and it was argued that de-labelling patients with penicillin allergy is unnecessary. This may suggest that in many cases, cefazolin is deemed safe for patients with a penicillin allergy.

3. A quality improvement initiative at a children's hospital sought to increase the use of cefazolin in patients with penicillin allergy labels undergoing orthopedic surgery. The initiative found that appropriate clarification and documentation of penicillin allergy increased the use of cefazolin for surgical prophylaxis from 50% to 74%. Furthermore, patients whose penicillin allergy was clarified were 4 times more likely to receive cefazolin.

4. A pharmacist-led project aimed at increasing the use of cefazolin in patients with a penicillin allergy label found that after implementation, β-lactam usage (including cefazolin) increased from 12.6% to 37.8%. The increase in cefazolin usage did not correlate with a significant increase in adverse reactions (0% vs 0.4%).

In conclusion, the texts suggest that cefazolin is frequently used and can be safe for patients with a history of penicillin allergy, especially when the penicillin allergy is clarified or when patients are appropriately risk stratified. However, a small number of patients may experience cross-reactivity between cefazolin and penicillin. Always, the full studies might contain more nuanced information or limitations.

**User**

Great, now with these abstracts, we are going to read the text and based on the content we will ask "is it safe to give cefazolin to a patient with a history of penicillin allergy?” - however we do not want external information that you may know - we only want to see what facts are present in the following text. We will also iterate through several chucks:

```

In our cohort, a pharmacy-led antibiotic selection algorithm for patients with a PAL receiving perioperative antimicrobial prophylaxis resulted in increased use of β-lactam antibiotics, decreased use of second-line antibiotics, and decreased costs without a significant change in the incidence of adverse reactions.

Perioperative cefazolin administration for total joint arthroplasty is a first-line antibiotic recommended by the American Academy of Orthopaedic Surgeons (AAOS) guidelines for the prevention of periprosthetic joint infections (PJIs). We aim to analyze the clinical viability of giving patients with a documented penicillin allergy (PA) a perioperative full-strength cefazolin "test dose" under anesthesia.

This is a retrospective chart review of 2,451 total joint arthroplasties from a high-volume arthroplasty orthopaedic surgeon over a 5-year period from January 2013 through December 2017. This surgeon routinely gave patients with a documented PA a full-strength cefazolin test dose while under anesthesia instead of administrating a second-line antibiotic. The primary outcomes examined were allergic reaction and postoperative infection.

Cefazolin was given to 87.1% of all patients (1,990) and 46.0% of patients with a PA (143). The total rate of allergic reactions among all patients was 0.5% (11). Only one patient with a documented PA who received cefazolin had an allergic reaction. The reaction was not severe and did not require any additional treatment. In patients who had no reported allergies and received cefazolin, 0.3% (6) had an allergic reaction. There was no statistically significant difference in the rate of allergic reaction when comparing patients with and without a PA (P = 0.95). Patients receiving cefazolin had an overall PJI rate of 2.9% (57) versus those patients receiving antibiotics other than cefazolin who sustained a 5.5% PJI rate (16), which was statistically significant (P = 0.02).

This study found that utilization of a full-strength test dose of cefazolin in patients with a documented PA is a feasible, safe, and effective way of increasing the rate of cefazolin administration and thus mitigating the risk of PJIs.

Emerging literature has detailed the safe use of cefazolin in patients with immunoglobulin E-mediated penicillin allergy labeling (PAL) such as hives and anaphylaxis. The purpose of this article is to detail efforts led by an antimicrobial stewardship pharmacist working with an interdisciplinary team to optimize preoperative antimicrobials in patients with PAL.

A pharmacist-led, interdisciplinary collaborative practice agreement (CPA) was activated in January 2020 to permit pharmacists to independently optimize preoperative antibiotics to the preferred cefazolin in patients with PAL if nonsevere or severe reactions had been reported. A patient registry was established covering the timeframe between January 8, 2020, and January 6, 2022. Reaction during surgery was assessed via 2-provider documentation, which included surgeon and anesthesiology staff documentation of any complications during the procedure related to a suspected allergic safety event. Utilization of cefazolin, clindamycin, and vancomycin for preoperative prophylaxis was monitored before and after implementation of the CPA.

During the stated timeframe, 10,182 procedures and/or surgeries were completed on 1,572 (15.4%) patients with PAL and 659 (41.9%) patients previously reporting at least one reaction categorized as a severe reaction, which was hives for 71.2% of these patients. Of the 659 patients with PAL reporting a severe reaction, 356 received a preoperative cephalosporin (cefazolin, 98.8%; ceftriaxone, 1.2%) and tolerated it without a reported safety event, including 52 patients with PAL previously reporting anaphylaxis. An increase in preferred preoperative antimicrobial prophylaxis utilization was noted (cefazolin: 86% to 96.3%, P < 0.001; 2019 to 2021) with reductions noted in the use of nonpreferred preoperative antibiotics (clindamycin: 2.1% to 0.2%, P < 0.001; vancomycin: 3.2% to 0.4%, P < 0.001; 2019 to 2021).

```

**ChatGPT**

Based on the provided texts, these are the facts concerning the question "is it safe to give cefazolin to a patient with a history of penicillin allergy?":

1. The first excerpt states that a pharmacy-led antibiotic selection algorithm for patients with penicillin allergy labels resulted in increased use of β-lactam antibiotics (such as cefazolin), decreased use of second-line antibiotics, and reduced costs, without a significant change in adverse reactions. This suggests that cefazolin use is safe and beneficial in patients with penicillin allergy labels.

2. The second study, which is a retrospective review of 2,451 total joint arthroplasties, showed that patients with a documented penicillin allergy (PA) were given a full-strength cefazolin "test dose" while under anesthesia. The total rate of allergic reactions among all patients was 0.5%, with only one allergic reaction from a patient with a documented PA who received cefazolin. This reaction wasn't severe and required no extra treatment. Furthermore, cefazolin use appeared to reduce the rate of postoperative infection. The study suggests that a full-strength test dose of cefazolin can be a feasible and safe option for patients with a documented PA.

3. In the third excerpt, an interdisciplinary practice agreement permitted pharmacists to independently optimize preoperative antibiotics to cefazolin in patients with penicillin allergy labels even if nonsevere or severe reactions had been reported. During the study period, a considerable number of patients with severe reported reactions received cefazolin and tolerated it without a reported safety event. This indicates that cefazolin is safe for use in patients with a history of penicillin allergy, including those who had previously reported severe reactions.

In conclusion, the provided texts support the safety of cefazolin use in patients with a history of penicillin allergy, with careful monitoring and correct management strategies in place. However, as always, the full studies might contain more nuanced information or limitations.

**User**

Great, now with these abstracts, we are going to read the text and based on the content we will ask "is it safe to give cefazolin to a patient with a history of penicillin allergy?” - however we do not want external information that you may know - we only want to see what facts are present in the following text. We will also iterate through several chucks:

```

A pharmacist-led, interdisciplinary CPA increased preferred preoperative antimicrobial use in patients with PAL reporting severe allergic reactions, including hives and anaphylaxis, without reported safety events.

To investigate the associations between intrapartum antibiotic prophylaxis of group B streptococcus (GBS) in pregnant women and the risk of food allergy in Chinese children.

Retrospective cohort study of 2,909 mother-child pairs.

Taixing People's Hospital in Eastern China.

Term infants born 2018-2019, followed longitudinally from birth to 3 years.

The GBS-IAP was defined as therapy with intravenous penicillin G or ampicillin or cefazolin ≥4 h prior to delivery to the mother. Reference infants were defined as born without or with other intrapartum antibiotic exposure.

To investigate the incidence information of food allergy in children aged 18 months and three years old. Kaplan-Meier survival analysis and log-rank tests were used to evaluate the cumulative incidence in the group with GBS-IAP and the group without GBS-IAP. Cox proportional hazards models were conducted to determine the univariate and multivariate association between maternal GBS-IAP and incident food allergy after various covariates were adjusted.

The cumulative incidence of food allergy in the group with GBS-IAP was higher than that in the group without GBS-IAP in children under 18 months old (8.1% vs. 4.5%, P = 0.005, log-rank test), but no significant differences were observed in children under three years old (9.2% vs. 7.0%, P = 0.146, log-rank test). The univariate cox proportional hazards model in children under 18 months old revealed that children in the GBS-IAP group had faster food allergy development when compared with children in the group without GBS-IAP (HR.: 1.887,95% CI: 1.207-2.950, P = 0.005), so was the multivariate model (HR.: 1.906,95% CI: 1.158-3.137, P = 0.011). However, both univariate (HR: 1.343, 95% CI: 0.891∼2.026, P = 0.159) and multivariate (HR: 1.253, 95%CI: 0.796∼1.972, P = 0.329) cox proportional hazards model in children under three years old showed no significant differences between children in the group with GBS-IAP and group without GBS-IAP.

Intrapartum antibiotic prophylaxis of group B streptococcus may increase the cumulative incidence and risk of food allergy in children under 18 months old, but it had no significant effect on children under three years old.

To understand the occurrence of atopic dermatitis (AD) in children aged 2 years on exposure to maternal group B streptococcus (GBS) antibiotic prophylaxis (IAP).

Retrospective cohort study of 2909 mother-child pairs.

Taixing People's Hospital in Eastern China.

Term infants born 2018-2019, followed longitudinally from birth to 2 years.

The GBS-IAP was defined as therapy with intravenous penicillin G or ampicillin or cefazolin ≥ 4 h prior to delivery to the mother. Reference infants were defined as born without or with other intrapartum antibiotic exposure.

The logistic regression models were employed to analyze the effect of intrapartum GBS prophylaxis on AD in 2-year-old children during delivery. Analysis was a priori stratified according to the mode of delivery and adjusted for relevant covariates.

The cohorts showed that preventive GBS-IAP was potentially associated with increased incidence of AD in children delivered vaginally according to logistic regression models before and after covariate-adjusted treatment (OR: 6.719,95% CI: 4.730-9.544,P < 0.001;aOR: 6.562,95% CI: 4.302-10.008, P < 0.001).

Prophylactic treatment of intrapartum GBS may raise the risk of AD in vaginally delivered children. These findings highlight the need to better understand the risk between childhood AD and current GBS-IAP intervention strategies.

```

**ChatGPT**

Based on the provided texts, these are the facts concerning the question "is it safe to give cefazolin to a patient with a history of penicillin allergy?":

1. The first excerpt indicates that a pharmacist-led, interdisciplinary collaborative practice agreement (CPA) increased the use of preferred preoperative antimicrobials (including cefazolin) in patients with Penicillin Allergy Labels (PALs) reporting severe allergic reactions, such as hives and anaphylaxis, without any reported safety events. This implies that cefazolin can be safely given to patients with severe penicillin allergies.

2. The second excerpt is a study on the association between intrapartum antibiotic prophylaxis of group B streptococcus (GBS) in pregnant women, which can include cefazolin, and the risk of food allergy in their offspring. It suggests that this type of antibiotic prophylaxis may increase the risk of food allergies in children under 18 months but not in children under three years old. However, this study does not directly address the safety of cefazolin in patients with penicillin allergies.

3. The third excerpt is another study investigating the occurrence of atopic dermatitis (AD) in children exposed to maternal GBS antibiotic prophylaxis, including cefazolin. It indicates that prophylactic treatment with such antibiotics may increase the risk of AD in vaginally delivered children. Similar to the previous study, it doesn't directly speak to the safety of cefazolin in patients with penicillin allergies.

In summary, the provided texts still indicate the safety of cefazolin use in patients with a history of penicillin allergy, under careful monitoring and proper management. The potential effects on offspring as described in the second and third excerpts introduce another dimension to consider when assessing overall safety but do not directly relate to the penicillin-allergic patients themselves.

**User**

Great, now with these abstracts, we are going to read the text and based on the content we will ask "is it safe to give cefazolin to a patient with a history of penicillin allergy?” - however we do not want external information that you may know - we only want to see what facts are present in the following text. We will also iterate through several chucks:

```

This study estimated prophylactic antibiotic usage rates for the prevention of early-onset invasive neonatal group B Streptococcus infection among patients with penicillin allergy. Undertreatment (no antibiotics, underuse of cefazolin, overuse of clindamycin inconsistent with resistance patterns) and overtreatment (vancomycin use) were common. Academic hospitals were marginally more adherent to guidelines than nonacademic hospitals.

Clinical risk factors for nephrotoxicity in Staphylococcus aureus bacteraemia remain largely undetermined, despite its common occurrence and clinical significance. In an international, multicentre, prospective clinical trial (CAMERA2), which compared standard therapy (vancomycin monotherapy) to combination therapy (adding an anti-staphylococcal beta-lactam) for methicillin-resistant S. aureus bacteraemia, significantly more people in the combination therapy arm experienced acute kidney injury compared with those in the monotherapy arm (23% vs 6%).

The aim of this post hoc analysis was to explore in greater depth the risk factors for acute kidney injury from the CAMERA2 trial.

Among participants of the CAMERA2 trial, demographic-related, infection-related and treatment-related risk factors were assessed for their relationship with acute kidney injury by univariable and multivariable logistic regression. Acute kidney injury was defined by a modified-KDIGO (Kidney Disease: Improving Global Outcomes) criteria (not including urinary output).

Of the 266 participants included, age (p = 0.04), randomisation to combination therapy (p = 0.002), vancomycin area under the concentration-time curve (p = 0.03) and receipt of (flu)cloxacillin as the companion beta-lactam (p < 0.001) were significantly associated with acute kidney injury. On a multivariable analysis, concurrent use of (flu)cloxacillin increased the risk of acute kidney injury over four times compared with the use of cefazolin or no beta-lactam. The association of vancomycin area under the concentration-time curve with acute kidney injury also persisted in the multivariable model.

For participants receiving vancomycin for S. aureus bacteraemia, use of (flu)cloxacillin and increased vancomycin area under the concentration-time curve were risk factors for acute kidney injury. These represent potentially modifiable risk factors for nephrotoxicity and highlight the importance of avoiding the use of concurrent nephrotoxins.

The use of alternative antibiotics to cefazolin has been associated with increased risk of infection after shoulder arthroplasty. Routine preoperative allergy testing in patients reporting penicillin and cephalosporin allergies increases the number able to receive cefazolin and may reduce the occurrence of infection after shoulder arthroplasty, but the financial viability of this practice is unclear. We used break-even modeling to determine the economic viability of routine preoperative allergy testing for infection prevention in total shoulder arthroplasty patients reporting penicillin and cephalosporin allergies.

Allergy testing cost ($248.24), infection rates after shoulder arthroplasty following use of noncefazolin antibiotics (3.09%), and infection-related care costs ($55,243) were derived from the literature. A break-even equation using these variables was developed to determine the absolute risk reduction (ARR) in the infection rate that would economically justify the routine implementation of preoperative allergy testing. The number needed to treat was calculated from the ARR.

Preoperative allergy testing is considered economically justified if it prevents at least 1 infection out of 223 shoulder arthroplasties (ARR = 0.45%). These protocols remained economically viable at varying allergy testing costs, initial infection rates, and infection-related care costs.

Routine preoperative penicillin allergy testing is an economically justified infection prevention strategy among patients reporting penicillin and cephalosporin allergies in the setting of elective shoulder arthroplasty. Widespread implementation of this practice may considerably reduce the economic and societal burden associated with prosthetic infections.

Cephalosporins are the cornerstone of cardiac device infection prophylaxis. Owing to fears of cross-reactivity, penicillin-allergic patients are exposed to potentially more-toxic drugs, with decreased efficacy. We evaluated the safety of a cefazolin test dose (CTD) in self-reported penicillin-allergic patients.

```

**ChatGPT**

Based on the provided texts, these are the facts concerning the question "is it safe to give cefazolin to a patient with a history of penicillin allergy?":

1. The first excerpt suggests that underuse of cefazolin is considered a form of undertreatment for the prevention of early-onset invasive neonatal group B Streptococcus infection in patients with penicillin allergy. This indicates that cefazolin is considered a safe and effective treatment option in these circumstances, which may indirectly suggest its safety in patients with penicillin allergies.

2. The second and third excerpts do not directly address the safety of cefazolin in patients with a history of penicillin allergy. However, they do suggest that the use of certain antibiotics like (flu)cloxacillin along with vancomycin can increase the risk of acute kidney injury. Cefazolin was compared favorably to (flu)cloxacillin, indicating a lower associated risk of kidney injury, which might imply a relative safety advantage for cefazolin in general.

3. The fourth excerpt suggests that the use of alternative antibiotics to cefazolin has been associated with an increased risk of infection after shoulder arthroplasty. It proposes that routine preoperative allergy testing in patients reporting penicillin and cephalosporin allergies might increase the number of patients able to receive cefazolin and potentially reduce the occurrence of infection after shoulder arthroplasty. This indirectly suggests the safety and preferability of cefazolin, even for patients with reported allergies.

4. The fifth excerpt directly mentions the evaluation of the safety of a cefazolin test dose in self-reported penicillin-allergic patients. This implies a clinical inquiry into the safety of cefazolin in penicillin-allergic patients.

In summary, while the provided texts do not explicitly answer the question, they suggest that cefazolin can be safely and effectively used in certain circumstances and may be preferable to other antibiotics. They also highlight the potential value of allergy testing for patients with a reported history of penicillin allergy, which could potentially increase the number of patients safely able to receive cefazolin.

**User**

Great, now with these abstracts, we are going to read the text and based on the content we will ask "is it safe to give cefazolin to a patient with a history of penicillin allergy?” - however we do not want external information that you may know - we only want to see what facts are present in the following text. We will also iterate through several chucks:

```

In this single-centre study, we evaluated consecutive patients with chart documentation of penicillin allergy undergoing cardiac device implantation, over a 2-year period. A CTD was performed if no cephalosporin allergy or severe anaphylactic reaction to penicillin had been documented. Patients were given 2 doses of 100 mg IV cefazolin, and if no allergic reaction occurred after 5 minutes, the full dose (1800 mg) was administered in the electrophysiology laboratory just before the implantation procedure.

A total of 2200 patients were included. The frequency of reported penicillin allergy was 9.3% (n = 204). In 80% of cases, the type of allergic reaction was not reported in medical notes or was unknown by the patient. A CTD was performed in 67.6% of patients with a penicillin allergy (n = 138). A total of 5 adverse events occurred (3.6% of patients [95% confidence interval, 1.1%-6.1%]) - 4 skin rashes and 1 tongue edema. These 5 patients became asymptomatic after antihistaminic and corticosteroid IV treatment. Even if the test dose was negative, 79% of patients also were administered vancomycin before the procedure, as it requires a 1-hour infusion prior to the CTD in the implantation procedure room.

A CTD in most penicillin-allergic patients appears to be safe and allows its use per recommended guidelines.

Les céphalosporines sont la pierre angulaire de la prophylaxie des infections des dispositifs cardiaques. En raison du risque appréhendé de réactivité croisée, les patients allergiques à la pénicilline se trouvent exposés à des médicaments potentiellement plus toxiques, qui s’avèrent aussi moins efficaces. Nous avons évalué l’innocuité d’une dose d’essai de céfazoline chez des patients qui s’étaient dits allergiques à la pénicilline.

Dans cette étude monocentrique, nous avons suivi pendant deux ans des patients consécutifs dont le dossier médical faisait état d’une allergie à la pénicilline et chez qui un dispositif cardiaque devait être implanté. Une dose d’essai de céfazoline a été administrée aux patients sans antécédents documentés d’allergie aux céphalosporines ou de réaction anaphylactique sévère à la pénicilline. Deux doses de 100 mg de céfazoline ont été administrées par voie intraveineuse. En l’absence de réaction allergique après cinq minutes, les patients recevaient la dose complète (1 800 mg) au laboratoire d’électrophysiologie juste avant l’implantation du dispositif cardiaque.

Au total, 2 200 patients ont été inscrits à l’étude. Le taux de signalement de l’allergie à la pénicilline était de 9,3 % (n = 204). Dans 80 % des cas, le type de réaction allergique n’a pas été précisé dans les notes médicales ou était inconnu du patient. Une dose d’essai de céfazoline a été administrée à 67,6 % des patients allergiques à la pénicilline (n = 138). Au total, cinq événements indésirables se sont produits (3,6 % des patients [intervalle de confiance à 95 % : 1,1-6,1 %]) – quatre éruptions cutanées et un œdème de la langue. Les cinq patients touchés par ces événements sont devenus asymptomatiques après avoir reçu un antihistaminique et un corticostéroïde par voie intraveineuse. Même en l’absence de réaction allergique à la dose d’essai, 79 % des patients ont reçu de la vancomycine avant l’intervention, cet agent devant être administré par perfusion durant une heure avant la dose d’essai de céfazoline dans la salle d’intervention.

Chez la plupart des patients allergiques à la pénicilline, une dose d’essai de céfazoline semble sans danger et permet d’avoir recours à ce médicament conformément aux lignes directrices.

Surgical site infections (SSI) can represent a major complication of pancreaticoduodenectomy (PD). We summarize the outcomes of process improvement efforts to reduce the SSI rates in PD that includes replacing Cefazolin with Ceftriaxone-Metronidazole as antibiotic prophylaxis. Additional efforts included current assessment of biliary microbiome and potential prophylactic failures based on bile cultures and suspected antibiotic allergies.

```

**ChatGPT**

Based on the provided texts, these are the facts concerning the question "is it safe to give cefazolin to a patient with a history of penicillin allergy?":

1. The first two excerpts, which appear to be the same study but in different languages (English and French), report on the safety of a cefazolin test dose in patients with a history of penicillin allergy. According to the study, in a group of 138 patients given a cefazolin test dose, only 5 adverse events were observed (four skin rashes and one case of tongue edema), representing 3.6% of patients. All of these patients became asymptomatic after treatment with an antihistamine and a corticosteroid. The study concludes that a cefazolin test dose in most penicillin-allergic patients appears to be safe.

2. The third excerpt does not directly address the safety of cefazolin in patients with a history of penicillin allergy. It discusses a change in antibiotic prophylaxis to reduce surgical site infection rates in pancreaticoduodenectomy, replacing Cefazolin with Ceftriaxone-Metronidazole. However, it doesn't provide any evidence relating to the safety or risk of cefazolin in patients with a history of penicillin allergy.

In summary, according to the results of the clinical study presented, it appears that cefazolin may be safely administered to most patients with a reported history of penicillin allergy, as long as no severe anaphylactic reaction to penicillin has been previously documented. However, a small percentage of patients might still experience allergic reactions, which were treatable with antihistamines and corticosteroids in this study.

**User**

Great, now with these abstracts, we are going to read the text and based on the content we will ask "is it safe to give cefazolin to a patient with a history of penicillin allergy?” - however we do not want external information that you may know - we only want to see what facts are present in the following text. We will also iterate through several chucks:

```

A single-center review of PD patients from January-2012 to March-2021. Study groups were divided into Pre and Post May-2015 (Group 1 and 2, respectively) when Ceftriaxone-Metronidazole prophylaxis and routine intraoperative cultures were standardized. Univariate and multivariable analyses were conducted to assess groups' differences and association with SSI.

Six hundred ninety patients identified [267(38.7%) and 423(61.3%) in Group 1 and Group2, respectively]. After antibiotic change, SSI rates decreased from 28.1% to 16.5% (incisional: 17.6%-7.5%, organ-space or abscess: 17.2%-13.0%), Group 1 and Group 2, respectively, P<0.001. Ceftriaxone-Metronidazole was used in 75.9% of patients Group 2. When adjusting for other covariates, an SSI-decrease was associated only with Ceftriaxone-Metronidazole (OR 0.34, P<0.001).

Ongoing process improvement has resulted in decreased SSIs with Ceftriaxone-Metronidazole prophylaxis. The benefit of Ceftriaxone-Metronidazole is independent of the biliary microbiome. Improving prophylaxis for those with suspected penicillin allergy is warranted.

Introduction  Improved evidence-based guidelines on the optimal type and duration of antibiotics for patients undergoing endoscopic endonasal transsphenoidal surgery (EETS) are needed. We analyze the infectious complications among a large cohort of EETS patients undergoing a standardized regimen of cefazolin for 24 hours, followed by cephalexin for 7 days after surgery (clindamycin if penicillin/cephalosporin allergic). Methods  A retrospective review of 132 EETS patients from 2018 to 2020 was conducted. Patient, tumor, and surgical characteristics were collected, along with infection rates. Multivariate logistic regression determined the variable(s) independently associated with infectious outcomes. Results  Nearly all patients (99%) received postoperative antibiotics with 78% receiving cefazolin, 17% receiving cephalexin, 3% receiving clindamycin, and 2% receiving other antibiotics. Fifty-three patients (40%) had an intraoperative cerebrospinal fluid (CSF) leak, and three patients (2%) developed a postoperative CSF leak requiring surgical repair. Within 30 days, no patients developed meningitis. Five patients (4%) developed sinusitis, two patients (3%) developed pneumonia, and one patient (1%) developed cellulitis at a peripheral intravenous line. Two patients (2%) developed an allergy to cephalexin, requiring conservative management. After adjustment for comorbidities and operative factors, presence of postoperative infectious complications was independently associated with increased LOS ( β  = 3.7 days; p  = 0.001). Conclusion  Compared with reported findings in the literature, we report low rates of infectious complications and antibiotic intolerance, despite presence of a heavy burden of comorbidities and high intraoperative CSF leak rates among our cohort. These findings support our standardized 7-day perioperative antibiotic regimen.

Clindamycin serves as an alternative surgical prophylactic antibiotic in patients with penicillin (PCN) or cephalosporin allergy labels. In the previous reports, the use of clindamycin was associated with higher incidences of surgical site infections (SSIs). We aimed to determine the characteristics of PCN or cephalosporin allergic reactions to stratify patient's risk and indicate subsequent management; leading to de-labeling of PCN or cephalosporin allergy.

We conducted a prospective cohort study of patients receiving clindamycin as surgical antibiotic prophylaxis from September 2021 to March 2022. Data were collected from electronic medical records; included demographic data, antibiotic allergy labels, allergic reaction, and allergy testing.

Clindamycin was administered in 445 patients who underwent 451 operations. Among these patients, 53.0% (n = 236) were female with a median age of 15 years (range; 0.5-57.0 years). PCN and cephalosporin allergies were labelled in 83.8% (n = 373) and 25.6% (n = 114) patients, respectively; 11.4% (n = 51) of patients were allergic to both classes of the antibiotics. There were 191 (51.2%) and 73 (64.0%) possible hypersensitivity reactions (HSRs) in PCN and cephalosporin groups, respectively. The most common reactions were rash (PCN: n = 99, 26.5%; cephalosporin: n = 35, 30.7%), and hives (PCN: n = 71, 19.0%; cephalosporin: n = 24, 21.1%). Severe reactions included angioedema (PCN: n = 7, 1.9%; cephalosporin: n = 5, 4.4%), anaphylaxis (PCN: n = 8, 2.1%; cephalosporin: n = 7, 6.1%), bronchospasm (cephalosporin: n = 1, 0.9%), airway involvement (PCN: n = 1, 0.3%; cephalosporin: n = 1, 0.9%), serum sickness (PCN: n = 1, 0.3%), blisters (PCN: n = 1, 0.3%), and drug reaction with eosinophilia and systemic symptoms (DRESS) (PCN: n = 1, 0.3%). Low-risk history of allergy included gastrointestinal side effects (PCN: n = 9, 2.4%; cephalosporin: n = 3, 2.7%), positive family history (PCN: n = 7, 1.9%; cephalosporin: n = 1, 0.9%), and remote history of allergy (PCN: n = 2, 0.5%). There were 201 (53.9%) and 53 (46.5%) unknown reactions in PCN and cephalosporin groups, respectively. In the overall cohort, 3 patients (0.7%) were skin tested for drug allergy (PCN: n = 2, 0.5%; cephalosporin: n = 2, 1.8%).

Clindamycin was largely administered in patients with non-severe HSRs, low-risk history or unknown reactions to PCN or cephalosporin, whom cefazolin could have been administered safely. Obtaining a detailed history of antibiotic allergy, allergy testing and/or direct oral challenge can de-label unsubstantiated PCN or cephalosporin allergy and ultimately reduce the incidence of SSIs by optimizing the rate of more effective antibiotic administration.

```

**ChatGPT**

Based on the information given in the text, here is what we can conclude in relation to the question "Is it safe to give cefazolin to a patient with a history of penicillin allergy?"

1. The text does not provide direct information on the safety of giving cefazolin to a patient with a history of penicillin allergy.

2. However, it does mention in the last paragraph that clindamycin, an alternative to cefazolin, was largely administered in patients with non-severe hypersensitivity reactions, low-risk history or unknown reactions to penicillin or cephalosporins, indicating that cefazolin could have been administered safely in these cases.

3. The text also emphasizes the need for obtaining a detailed history of antibiotic allergy, allergy testing, and/or direct oral challenge to de-label unsubstantiated penicillin or cephalosporin allergy. This suggests that with proper medical screening, it could be determined whether it would be safe to administer cefazolin to a patient with a history of penicillin allergy.

However, these conclusions do not come from direct testing or experimentation with cefazolin in penicillin-allergic patients according to the provided text. As such, further research or information would be needed to directly answer the posed question based solely on this text.

**User**

The following text contains summaries of 90 abstracts, we are going to read the text and based on the content we will ask "is it safe to give cefazolin to a patient with a history of penicillin allergy?” - however we do not want external information that you may know - we only want to see what facts are present in the following text.

I will provide the content in two parts - please do not answer until you have PART 1 and PART 2.

PART 1 of 2:

```

Based on the given abstracts, here are the relevant points that can help answer the question "Is it safe to give cefazolin to a patient with a history of penicillin allergy?"

1. In a retrospective review involving patients labeled allergic to penicillin or cephalosporin antibiotics who underwent a primary total hip and/or knee arthroplasty, there were fewer surgical site infections (SSIs) in the cefazolin group compared with the clindamycin and/or vancomycin group. Importantly, the frequency of intraoperative hypersensitivity reactions (HSRs) was not significantly different between the groups (cefazolin = 0.2% vs clindamycin/vancomycin = 1.3%; P = .06).

Conclusion: The use of cefazolin as a perioperative antibiotic in these surgical procedures in patients labeled as beta-lactam allergic was associated with a decreased rate of postoperative SSIs without an increase in intraoperative HSR.

2. In a retrospective cohort study of Group B Streptococcus (GBS) prophylaxis in pregnant individuals, 259 out of 1170 (22.1%) mothers with a documented penicillin allergy received cefazolin. The study showed that neonates of patients who received alternative GBS prophylaxis (vancomycin or clindamycin) were significantly more likely to undergo a postnatal lab draw compared to neonates of patients who received beta-lactam antibiotics (including cefazolin), but there were no other significant differences in newborn outcomes.

Conclusion: Cefazolin was administered to a significant proportion of mothers with a documented penicillin allergy for GBS prophylaxis during pregnancy, and there were no mentioned adverse outcomes specific to this group.

Based on these abstracts, cefazolin was administered to patients with a history of penicillin allergy in different scenarios and did not demonstrate an increase in hypersensitivity reactions or other adverse outcomes compared to alternative antibiotics. However, these are abstracts and the full studies might contain more nuanced information or limitations.

Based on the given abstracts, here are the relevant points that can help answer the question "Is it safe to give cefazolin to a patient with a history of penicillin allergy?"

1. A study of pregnant individuals who reported a penicillin allergy revealed that they were more likely to receive alternative antibiotics for GBS prophylaxis, which led to an increased frequency of postnatal blood draws for their neonates. However, the abstract does not specifically discuss outcomes related to the administration of cefazolin in these cases.

2. The Allergy Clarification for Cefazolin Evidence-based Prescribing Tool (ACCEPT) was developed and implemented to guide the use of perioperative cefazolin in patients with reported beta-lactam allergies undergoing cesarean delivery. Of the eligible women, 9% reported a beta-lactam allergy, with penicillin being the most common allergen. The use of cefazolin increased significantly from 52% (baseline) to 87% during the intervention period. Notably, there was only a small increase in perioperative allergic reactions (1 in the baseline period and 2 during the intervention period), suggesting that the administration of cefazolin was generally safe.

3. The implementation of a guideline identifying when inpatients with penicillin or cephalosporin antibiotic allergy labels (PCAAL) can receive β-lactam antibiotics led to an increase in β-lactam receipt. In the Medical-PCAAL group, the use of cefazolin specifically increased (from 8% to 11%), and there were no significant changes in these outcomes on surgical or obstetrics/gynecology services. The abstract does not mention any increase in allergic reactions or adverse outcomes associated with this increase in cefazolin use.

Based on these abstracts, it appears that in certain clinical situations, cefazolin was safely administered to patients with a history of penicillin allergy without a significant increase in allergic reactions or adverse outcomes. However, these are abstracts and the full studies might contain more nuanced information or limitations.

Based on the given abstracts, here are the relevant points that can help answer the question "Is it safe to give cefazolin to a patient with a history of penicillin allergy?"

1. A retrospective study aimed at determining which antibiotics were used for GBS positive patients with documented penicillin allergies found that cefazolin was frequently prescribed for intrapartum neonatal sepsis prophylaxis. While the study did not provide specific safety outcomes for the patients who received cefazolin, it indicated that the antibiotic choice followed the current guidelines of the American College of Obstetricians and Gynecologists (ACOG).

2. In relation to shoulder surgery infection prophylaxis, it was mentioned that IV cefazolin has been shown to be more effective than antibiotic alternatives such as vancomycin. Further, the text recommends that patients with a questionable history of penicillin allergy should undergo additional testing, potentially suggesting that cefazolin may be safe to use in some patients with a history of penicillin allergy, following further allergenic testing.

3. Cephalosporins, such as cefazolin, are presented as the preferred antibiotics for prophylaxis against surgical site infections. However, it is recommended that cephalosporins be avoided in patients with a history of IgE penicillin allergy but can be used in those with non-IgE penicillin allergy.

In conclusion, based on the given abstracts, it appears that cefazolin is frequently used and potentially safe for patients with a history of penicillin allergy, especially in cases with non-IgE penicillin allergy or those with a questionable history of allergy who undergo additional testing. As always, these are abstracts and the full studies might contain more nuanced information or limitations.

```

END OF PART 1 of 2...

**ChatGPT**

Acknowledged. I am waiting for PART 2 of 2.

**User**

PART 2 of 2:

```

Based on the text provided, the following facts can be established regarding the safety of giving cefazolin to a patient with a history of penicillin allergy:

1. The cross-reactivity rate between cefazolin and both non-IgE and IgE reactions to penicillin was relatively low, at 0.9% and 4.0% respectively. Similar rates were observed for those who reported reactions to the penicillin family. The rates of cross reactivity were found to be lower than previously reported in older literature but consistent with more recent literature.

2. A report indicates that cefazolin is frequently the preferred antibiotic for prophylaxis against surgical site infections, and it was argued that de-labelling patients with penicillin allergy is unnecessary. This may suggest that in many cases, cefazolin is deemed safe for patients with a penicillin allergy.

3. A quality improvement initiative at a children's hospital sought to increase the use of cefazolin in patients with penicillin allergy labels undergoing orthopedic surgery. The initiative found that appropriate clarification and documentation of penicillin allergy increased the use of cefazolin for surgical prophylaxis from 50% to 74%. Furthermore, patients whose penicillin allergy was clarified were 4 times more likely to receive cefazolin.

4. A pharmacist-led project aimed at increasing the use of cefazolin in patients with a penicillin allergy label found that after implementation, β-lactam usage (including cefazolin) increased from 12.6% to 37.8%. The increase in cefazolin usage did not correlate with a significant increase in adverse reactions (0% vs 0.4%).

In conclusion, the texts suggest that cefazolin is frequently used and can be safe for patients with a history of penicillin allergy, especially when the penicillin allergy is clarified or when patients are appropriately risk stratified. However, a small number of patients may experience cross-reactivity between cefazolin and penicillin. Always, the full studies might contain more nuanced information or limitations.

Based on the provided texts, these are the facts concerning the question "is it safe to give cefazolin to a patient with a history of penicillin allergy?":

1. The first excerpt states that a pharmacy-led antibiotic selection algorithm for patients with penicillin allergy labels resulted in increased use of β-lactam antibiotics (such as cefazolin), decreased use of second-line antibiotics, and reduced costs, without a significant change in adverse reactions. This suggests that cefazolin use is safe and beneficial in patients with penicillin allergy labels.

2. The second study, which is a retrospective review of 2,451 total joint arthroplasties, showed that patients with a documented penicillin allergy (PA) were given a full-strength cefazolin "test dose" while under anesthesia. The total rate of allergic reactions among all patients was 0.5%, with only one allergic reaction from a patient with a documented PA who received cefazolin. This reaction wasn't severe and required no extra treatment. Furthermore, cefazolin use appeared to reduce the rate of postoperative infection. The study suggests that a full-strength test dose of cefazolin can be a feasible and safe option for patients with a documented PA.

3. In the third excerpt, an interdisciplinary practice agreement permitted pharmacists to independently optimize preoperative antibiotics to cefazolin in patients with penicillin allergy labels even if nonsevere or severe reactions had been reported. During the study period, a considerable number of patients with severe reported reactions received cefazolin and tolerated it without a reported safety event. This indicates that cefazolin is safe for use in patients with a history of penicillin allergy, including those who had previously reported severe reactions.

In conclusion, the provided texts support the safety of cefazolin use in patients with a history of penicillin allergy, with careful monitoring and correct management strategies in place. However, as always, the full studies might contain more nuanced information or limitations.

Based on the provided texts, these are the facts concerning the question "is it safe to give cefazolin to a patient with a history of penicillin allergy?":

1. The first excerpt indicates that a pharmacist-led, interdisciplinary collaborative practice agreement (CPA) increased the use of preferred preoperative antimicrobials (including cefazolin) in patients with Penicillin Allergy Labels (PALs) reporting severe allergic reactions, such as hives and anaphylaxis, without any reported safety events. This implies that cefazolin can be safely given to patients with severe penicillin allergies.

2. The second excerpt is a study on the association between intrapartum antibiotic prophylaxis of group B streptococcus (GBS) in pregnant women, which can include cefazolin, and the risk of food allergy in their offspring. It suggests that this type of antibiotic prophylaxis may increase the risk of food allergies in children under 18 months but not in children under three years old. However, this study does not directly address the safety of cefazolin in patients with penicillin allergies.

3. The third excerpt is another study investigating the occurrence of atopic dermatitis (AD) in children exposed to maternal GBS antibiotic prophylaxis, including cefazolin. It indicates that prophylactic treatment with such antibiotics may increase the risk of AD in vaginally delivered children. Similar to the previous study, it doesn't directly speak to the safety of cefazolin in patients with penicillin allergies.

In summary, the provided texts still indicate the safety of cefazolin use in patients with a history of penicillin allergy, under careful monitoring and proper management. The potential effects on offspring as described in the second and third excerpts introduce another dimension to consider when assessing overall safety but do not directly relate to the penicillin-allergic patients themselves.

Based on the provided texts, these are the facts concerning the question "is it safe to give cefazolin to a patient with a history of penicillin allergy?":

1. The first excerpt suggests that underuse of cefazolin is considered a form of undertreatment for the prevention of early-onset invasive neonatal group B Streptococcus infection in patients with penicillin allergy. This indicates that cefazolin is considered a safe and effective treatment option in these circumstances, which may indirectly suggest its safety in patients with penicillin allergies.

2. The second and third excerpts do not directly address the safety of cefazolin in patients with a history of penicillin allergy. However, they do suggest that the use of certain antibiotics like (flu)cloxacillin along with vancomycin can increase the risk of acute kidney injury. Cefazolin was compared favorably to (flu)cloxacillin, indicating a lower associated risk of kidney injury, which might imply a relative safety advantage for cefazolin in general.

3. The fourth excerpt suggests that the use of alternative antibiotics to cefazolin has been associated with an increased risk of infection after shoulder arthroplasty. It proposes that routine preoperative allergy testing in patients reporting penicillin and cephalosporin allergies might increase the number of patients able to receive cefazolin and potentially reduce the occurrence of infection after shoulder arthroplasty. This indirectly suggests the safety and preferability of cefazolin, even for patients with reported allergies.

4. The fifth excerpt directly mentions the evaluation of the safety of a cefazolin test dose in self-reported penicillin-allergic patients. This implies a clinical inquiry into the safety of cefazolin in penicillin-allergic patients.

In summary, while the provided texts do not explicitly answer the question, they suggest that cefazolin can be safely and effectively used in certain circumstances and may be preferable to other antibiotics. They also highlight the potential value of allergy testing for patients with a reported history of penicillin allergy, which could potentially increase the number of patients safely able to receive cefazolin.

Based on the provided texts, these are the facts concerning the question "is it safe to give cefazolin to a patient with a history of penicillin allergy?":

1. The first two excerpts, which appear to be the same study but in different languages (English and French), report on the safety of a cefazolin test dose in patients with a history of penicillin allergy. According to the study, in a group of 138 patients given a cefazolin test dose, only 5 adverse events were observed (four skin rashes and one case of tongue edema), representing 3.6% of patients. All of these patients became asymptomatic after treatment with an antihistamine and a corticosteroid. The study concludes that a cefazolin test dose in most penicillin-allergic patients appears to be safe.

2. The third excerpt does not directly address the safety of cefazolin in patients with a history of penicillin allergy. It discusses a change in antibiotic prophylaxis to reduce surgical site infection rates in pancreaticoduodenectomy, replacing Cefazolin with Ceftriaxone-Metronidazole. However, it doesn't provide any evidence relating to the safety or risk of cefazolin in patients with a history of penicillin allergy.

In summary, according to the results of the clinical study presented, it appears that cefazolin may be safely administered to most patients with a reported history of penicillin allergy, as long as no severe anaphylactic reaction to penicillin has been previously documented. However, a small percentage of patients might still experience allergic reactions, which were treatable with antihistamines and corticosteroids in this study.

Based on the information given in the text, here is what we can conclude in relation to the question "Is it safe to give cefazolin to a patient with a history of penicillin allergy?"

1. The text does not provide direct information on the safety of giving cefazolin to a patient with a history of penicillin allergy.

2. However, it does mention in the last paragraph that clindamycin, an alternative to cefazolin, was largely administered in patients with non-severe hypersensitivity reactions, low-risk history or unknown reactions to penicillin or cephalosporins, indicating that cefazolin could have been administered safely in these cases.

3. The text also emphasizes the need for obtaining a detailed history of antibiotic allergy, allergy testing, and/or direct oral challenge to de-label unsubstantiated penicillin or cephalosporin allergy. This suggests that with proper medical screening, it could be determined whether it would be safe to administer cefazolin to a patient with a history of penicillin allergy.

However, these conclusions do not come from direct testing or experimentation with cefazolin in penicillin-allergic patients according to the provided text. As such, further research or information would be needed to directly answer the posed question based solely on this text.

```

**ChatGPT**

Based on the provided text, it seems that the issue of administering cefazolin to patients with a history of penicillin allergy is approached carefully in clinical settings. The collected data suggests the following:

1. The use of cefazolin among patients with reported allergies to penicillin tends to result in a low rate of cross-reactivity. This means that most patients with penicillin allergies don't experience allergic reactions when given cefazolin. However, a small percentage of patients may still have reactions due to cross-reactivity between penicillin and cefazolin.

2. Some studies indicate that de-labeling of patients with a penicillin allergy may not be necessary before administering cefazolin, suggesting a growing confidence in its safety profile among patients with a history of penicillin allergy.

3. Hospitals and pharmacies have successfully increased the use of cefazolin among penicillin-allergic patients, often without a corresponding increase in adverse reactions. This increase is made possible through initiatives that improve the understanding and documentation of the penicillin allergy, allowing healthcare professionals to make informed decisions.

4. Other texts suggest that underuse of cefazolin in certain situations is considered undertreatment, which implies a strong confidence in its effectiveness and safety profile. Other antibiotics may present additional risks compared to cefazolin, emphasizing its relative safety.

In summary, the provided texts suggest that with careful risk stratification, allergy clarification, and careful monitoring, cefazolin can be safely administered to patients with a history of penicillin allergy. However, the risk of cross-reactivity, though small, is still present and should always be taken into account. Therefore, the decision to use cefazolin should always be taken by a healthcare professional who has thoroughly evaluated the individual patient's allergy history and the benefits versus potential risks of the treatment.

**User**

Excellent, thank you very much and well done. Now I want to return to the R code and ```all_abstracts``` where we have source 90 abstracts from PubMed. We want to make a plot to illustrate some information about the question is it safe to give cefazolin to a patient with a history of penicillin allergy?” by using term-frequency to see what common terms appear in the dataset and then produce a ggplot to accompany the statement that we have generated from the same dataset.

**ChatGPT**

Certainly, to create a term-frequency plot, you will first need to extract the words from the abstracts and then count the frequency of each. Here's a simple example of how you can do this in R:

1. Install and load the necessary packages:

```R

install.packages(c("tidyverse", "tidytext", "ggplot2", "stopwords"))

library(tidyverse)

library(tidytext)

library(ggplot2)

library(stopwords)

```

2. Tokenize the abstracts, which means to break the text into individual words:

```R

tokenized_abstracts <- all_abstracts %>%

unnest_tokens(word, abstract)

```

3. Remove common "stop words" (like "and", "the", "of", etc.) as they are usually not informative:

```R

data(stop_words)

tokenized_abstracts <- tokenized_abstracts %>%

anti_join(stop_words)

```

4. Count the frequency of each word:

```R

word_counts <- tokenized_abstracts %>%

count(word, sort = TRUE)

```

5. Make the plot, showing the most common words:

```R

word_counts %>%

filter(n > quantile(n, 0.9)) %>% # Optional: Only show the most common words

ggplot(aes(reorder(word, n), n)) +

geom_col() +

coord_flip() +

labs(x = "Word",

y = "Frequency",

title = "Most Common Words in Abstracts Related to Cefazolin and Penicillin Allergy")

```

Please note that this is a basic example. Depending on your specific needs and the nature of your texts, you might want to preprocess your texts differently (e.g., remove punctuation, convert to lower case, etc.), use different tokenization techniques (e.g., n-grams), or visualize the data in different ways.

Remember to interpret the visualization with caution. High frequency of a word does not necessarily mean high importance, and the context in which a word is used is also very important. This visualization can, however, give you a broad overview of the main topics covered in the abstracts.

**User**

> tokenized_abstracts <- all_abstracts %>%

+	unnest_tokens(word, abstract)

Error in UseMethod("pull") :

no applicable method for 'pull' applied to an object of class "character"

**ChatGPT**

My apologies for the confusion. The error you encountered means that the ```all_abstracts``` object is not recognized as a dataframe by the ```pull``` function, which is used internally by ```unnest_tokens```. ```unnest_tokens``` is expecting a dataframe or a tibble.

If ```all_abstracts``` is a character vector where each element is an abstract, you would need to convert it to a tibble (or dataframe) first.

Here's how you can modify the code:

```r

\# Create a tibble

abstracts_tibble <- tibble(abstract = all_abstracts)

\# Tokenize the abstracts

tokenized_abstracts <- abstracts_tibble %>%

unnest_tokens(word, abstract)

\# Then you can proceed with the rest of the steps as before.

```

Try using this code, and let me know if you encounter any further issues!

**User**

excellent, here is the full code block. I now want to highlight our terms of interest on the plot - can we add color and geom_text to the terms defined earlier ```terms <- c("cefazolin", "penicillin", "allergy")``` ```library(tidyverse)

library(tidytext)

library(ggplot2)

library(stopwords)

\# Create a tibble

abstracts_tibble <- tibble(abstract = all_abstracts)

\# Tokenize the abstracts, which means to break the text into individual words:

tokenized_abstracts <- abstracts_tibble %>%

unnest_tokens(word, abstract)

\# Then you can proceed with the rest of the steps as before.

\# Remove common "stop words" (like "and", "the", "of", etc.) as they are usually not informative:

data(stop_words)

tokenized_abstracts <- tokenized_abstracts %>%

anti_join(stop_words)

\# Count the frequency of each word:

word_counts <- tokenized_abstracts %>%

count(word, sort = TRUE)

\# Make the plot, showing the most common words:

word_counts %>%

filter(n > quantile(n, 0.9)) %>% # Optional: Only show the most common words

ggplot(aes(reorder(word, n), n)) +

geom_col() +

coord_flip() +

labs(x = "Word",

y = "Frequency",

title = "Most Common Words in Abstracts Related to Cefazolin and Penicillin Allergy")

```

**ChatGPT**

Sure! Here's how you can add colors and labels to the terms of interest:

```r

\# Define the terms of interest

terms <- c("cefazolin", "penicillin", "allergy")

\# Make the plot, showing the most common words:

word_counts %>%

mutate(TermOfInterest = ifelse(word %in% terms, "Yes", "No")) %>%

filter(n > quantile(n, 0.9)) %>% # Optional: Only show the most common words

ggplot(aes(reorder(word, n), n, fill = TermOfInterest)) +

geom_col() +

geom_text(aes(label = ifelse(TermOfInterest == "Yes", as.character(word), "")), hjust = -0.1) +

scale_fill_manual(values = c("Yes" = "red", "No" = "grey50")) +

coord_flip() +

labs(x = "Word",

y = "Frequency",

title = "Most Common Words in Abstracts Related to Cefazolin and Penicillin Allergy",

fill = "Term of Interest")

```

In this code:

1. We create a new column in ```word_counts``` called ```TermOfInterest``` that is "Yes" for the words of interest and "No" otherwise.

2. We set the fill color of the bars in the plot to depend on ```TermOfInterest```.

3. We add text labels for the words of interest using ```geom_text```. The labels are positioned to the right of the bars (because of ```coord_flip```), and they only appear for the words of interest (the labels for the other words are empty strings).

4. We set the fill colors manually to be red for the words of interest and grey for the other words.

5. We add a legend title for the fill color.

**User**

great, lets save it as pdf using ggsave

**ChatGPT**

Absolutely. Here is how you can save the plot as a PDF using ```ggsave()```:

```r

\# Store your plot in a variable

plot <- word_counts %>%

mutate(TermOfInterest = ifelse(word %in% terms, "Yes", "No")) %>%

filter(n > quantile(n, 0.9)) %>% # Optional: Only show the most common words

ggplot(aes(reorder(word, n), n, fill = TermOfInterest)) +

geom_col() +

geom_text(aes(label = ifelse(TermOfInterest == "Yes", as.character(word), "")), hjust = -0.1) +

scale_fill_manual(values = c("Yes" = "red", "No" = "grey50")) +

coord_flip() +

labs(x = "Word",

y = "Frequency",

title = "Most Common Words in Abstracts Related to Cefazolin and Penicillin Allergy",

fill = "Term of Interest")

\# Save it as a PDF

ggsave(filename = "abstracts_word_frequency.pdf", plot = plot, width = 10, height = 7, units = "in")

```

In the ```ggsave``` function:

- ```filename``` is the name of the output file.

- ```plot``` is the plot to save (here, the last plot you created).

- ```width```, ```height```, and ```units``` specify the size of the output image. Adjust these values to suit your needs. You can specify the size in various units (like "cm", "mm", "in", "px", etc.), but here we are using inches ("in").

By default, ```ggsave``` will save the file to your current working directory. If you want to save it to a specific location, just include the full path in the ```filename``` argument (like ```"/path/to/your/directory/abstracts_word_frequency.pdf"```).

**User**

great, is there any other plot that would be good in this type of report where we used 90 abstracts on pubmed to summarise knowledge in the field?

**ChatGPT**

There are several types of visualizations that might be useful in this context:

1. **Word Cloud**: A word cloud is a popular way to visually represent text data. It can show the frequency of words in the abstracts as the size of the words.

2. **Correlation Plot or Network Analysis**: This could help show which words tend to appear together. This might be useful if, for example, you want to show which terms related to cefazolin and penicillin allergy often co-occur.

3. **Term Frequency-Inverse Document Frequency (TF-IDF) Plot**: While the term frequency plot we created above is useful, it might be dominated by somewhat common (but not necessarily meaningful) words. A TF-IDF plot can help identify words that are particularly characteristic of your abstracts, because it balances term frequency with the frequency of the term in a larger corpus (in this case, the whole set of abstracts).

4. **Sentiment Analysis**: Although this might be a bit more advanced, you could attempt to analyze the "sentiment" of the abstracts (e.g., are they positive/negative about the use of cefazolin in patients with a history of penicillin allergy?). This would require a sentiment lexicon, which is a list of words each associated with a sentiment score. Note that this is a somewhat subjective and complex process, and results should be interpreted carefully.

Which one do you think is most interesting or useful to you?

**User**

lets make a Term Frequency-Inverse Document Frequency (TF-IDF) Plot using ggplot and other tools. Note that we have already done ```

\# Create a tibble

abstracts_tibble <- tibble(abstract = all_abstracts)

\# Tokenize the abstracts, which means to break the text into individual words:

tokenized_abstracts <- abstracts_tibble %>%

unnest_tokens(word, abstract)

\# Then you can proceed with the rest of the steps as before.

\# Remove common "stop words" (like "and", "the", "of", etc.) as they are usually not informative:

data(stop_words)

tokenized_abstracts <- tokenized_abstracts %>%

anti_join(stop_words)

\# Count the frequency of each word:

word_counts <- tokenized_abstracts %>%

count(word, sort = TRUE)

\# Make the plot, showing the most common words:

word_counts %>%

filter(n > quantile(n, 0.9)) %>% # Optional: Only show the most common words

ggplot(aes(reorder(word, n), n)) +

geom_col() +

coord_flip() +

labs(x = "Word",

y = "Frequency",

title = "Most Common Words in Abstracts Related to Cefazolin and Penicillin Allergy")

\# Define the terms of interest

terms <- c("cefazolin", "penicillin", "allergy")

```

**ChatGPT**

Sure, here's how you could calculate and plot the Term Frequency-Inverse Document Frequency (TF-IDF) of your tokenized abstracts. The TF-IDF is a measure used to reflect how important a word is to a document in a collection or corpus. The importance increases proportionally to the number of times a word appears in the document and is offset by the number of documents in the corpus that contain the word.

```R

\# Calculate TF-IDF:

tf_idf <- tokenized_abstracts %>%

count(word) %>%

bind_tf_idf(word, n) %>%

arrange(desc(tf_idf))

\# Make the plot, showing the words with the highest TF-IDF:

tf_idf %>%

filter(n > quantile(n, 0.9)) %>% # Optional: Only show the most important words

ggplot(aes(reorder(word, tf_idf), tf_idf)) +

geom_col() +

coord_flip() +

labs(x = "Word",

y = "TF-IDF",

title = "Words with Highest TF-IDF in Abstracts Related to Cefazolin and Penicillin Allergy")

\# Highlight the terms of interest

terms <- c("cefazolin", "penicillin", "allergy")

tf_idf %>%

filter(word %in% terms) %>%

ggplot(aes(reorder(word, tf_idf), tf_idf)) +

geom_col(fill = "skyblue") +

coord_flip() +

labs(x = "Word",

y = "TF-IDF",

title = "TF-IDF for Terms of Interest in Abstracts Related to Cefazolin and Penicillin Allergy")

```

Please note, we are considering the term frequency within the abstract and the inverse document frequency across all abstracts. The 'term' in this case is considered as a word from the ```tokenized_abstracts```. The ```bind_tf_idf``` function from ```tidytext``` is used to calculate the TF-IDF. This function requires three arguments, a term, a document, and a count. Here, the 'document' corresponds to the abstracts.

This code will create two plots - one for all words and another just for your terms of interest. In both plots, the words are sorted in decreasing order of their TF-IDF. This should give you an idea of which words are particularly characteristic of your abstracts.

**User**

> # TF-IDF ----

> # Calculate TF-IDF:

> tf_idf <- tokenized_abstracts %>%

+	count(word) %>%

+	bind_tf_idf(word, n) %>%

+	arrange(desc(tf_idf))

Error in tapply(n, documents, sum) : arguments must have same length

**ChatGPT**

Apologies for the confusion. The error is due to incorrect usage of the ```bind_tf_idf()``` function. It requires three parameters: term, document, and count. The term is the individual word, count is the number of times it appears, and the document is the individual abstracts.

In this case, we need to first add an identifier for each individual abstract in your dataset before proceeding with the count. This will allow us to calculate the term frequency in each document (abstract) and the inverse document frequency across all documents (abstracts).

Here's how you can do this:

```R

\# Add a unique identifier for each abstract:

abstracts_tibble <- abstracts_tibble %>%

mutate(abstract_id = row_number())

\# Tokenize the abstracts again with the unique identifier:

tokenized_abstracts <- abstracts_tibble %>%

unnest_tokens(word, abstract)

\# Remove common "stop words" (like "and", "the", "of", etc.) as they are usually not informative:

data(stop_words)

tokenized_abstracts <- tokenized_abstracts %>%

anti_join(stop_words)

\# Now you can calculate TF-IDF:

tf_idf <- tokenized_abstracts %>%

count(abstract_id, word) %>%

bind_tf_idf(word, abstract_id, n) %>%

arrange(desc(tf_idf))

\# Plotting the words with highest TF-IDF value

tf_idf %>%

filter(n > quantile(n, 0.9)) %>% # Optional: Only show the most common words

ggplot(aes(reorder(word, tf_idf), tf_idf)) +

geom_col() +

coord_flip() +

labs(x = "Word",

y = "TF-IDF",

title = "Words with Highest TF-IDF in Abstracts Related to Cefazolin and Penicillin Allergy")

\# Highlighting the terms of interest

terms <- c("cefazolin", "penicillin", "allergy")

tf_idf %>%

filter(word %in% terms) %>%

ggplot(aes(reorder(word, tf_idf), tf_idf)) +

geom_col(fill = "skyblue") +

coord_flip() +

labs(x = "Word",

y = "TF-IDF",

title = "TF-IDF for Terms of Interest in Abstracts Related to Cefazolin and Penicillin Allergy")

```

This updated code first adds a unique identifier to each abstract. The ```unnest_tokens()``` function is then applied on the updated abstract tibble. Subsequently, we calculate the TF-IDF. Finally, two plots are created, one for all words and one for the terms of interest, highlighting them.

**User**

not that the result of this is only 1 row - perhaps we need to split it first, since we started with ```

\# Create the query string by joining the terms with '[Abstract] AND '

query <- paste0(terms, "[Abstract]", collapse = " AND ")

\# Search for publications in PubMed

papers <- entrez_search(db="pubmed", term=query)

\# Fetch records in "abstract" format

xmlData <- entrez_fetch(db="pubmed", id=papers$ids, rettype="abstract", retmode="xml")

\# Parse XML

doc <- xmlParse(xmlData)

\# Extract abstract nodes

abstractNodes <- getNodeSet(doc, "//AbstractText")

\# Extract the content of abstract nodes

abstracts <- sapply(abstractNodes, xmlValue)

\# Combine all abstracts into a single string, separated by two newline characters

all_abstracts <- paste(abstracts, collapse = "\n\n")

```

**ChatGPT**

Given that ```all_abstracts``` is a single string containing all the abstracts, you're right, we need to split it first into individual abstracts to treat each abstract as a document for the TF-IDF calculation.

Let's first split the ```all_abstracts``` into individual abstracts and then apply the TF-IDF:

```R

\# Split the single string of all_abstracts into individual abstracts

abstracts_tibble <- tibble(abstract = strsplit(all_abstracts, "\n\n")[[1]])

\# Add a unique identifier for each abstract:

abstracts_tibble <- abstracts_tibble %>%

mutate(abstract_id = row_number())

\# Tokenize the abstracts with the unique identifier:

tokenized_abstracts <- abstracts_tibble %>%

unnest_tokens(word, abstract)

\# Remove common "stop words" (like "and", "the", "of", etc.) as they are usually not informative:

data(stop_words)

tokenized_abstracts <- tokenized_abstracts %>%

anti_join(stop_words)

\# Calculate TF-IDF:

tf_idf <- tokenized_abstracts %>%

count(abstract_id, word) %>%

bind_tf_idf(word, abstract_id, n) %>%

arrange(desc(tf_idf))

\# Plotting the words with highest TF-IDF value

tf_idf %>%

filter(tf_idf > quantile(tf_idf, 0.9)) %>% # Optional: Only show words with highest TF-IDF values

ggplot(aes(reorder(word, tf_idf), tf_idf)) +

geom_col() +

coord_flip() +

labs(x = "Word",

y = "TF-IDF",

title = "Words with Highest TF-IDF in Abstracts Related to Cefazolin and Penicillin Allergy")

\# Highlighting the terms of interest

terms <- c("cefazolin", "penicillin", "allergy")

tf_idf %>%

filter(word %in% terms) %>%

ggplot(aes(reorder(word, tf_idf), tf_idf)) +

geom_col(fill = "skyblue") +

coord_flip() +

labs(x = "Word",

y = "TF-IDF",

title = "TF-IDF for Terms of Interest in Abstracts Related to Cefazolin and Penicillin Allergy")

```

By splitting the abstracts first, each abstract is treated as a document. This will allow for more meaningful TF-IDF scores. The code above should allow you to visualize words in terms of their importance across abstracts.

**User**

lets instead keep the top ranked terms with ```tf_idf %>%

\# filter(word %in% terms) %>%

filter(tf_idf > quantile(tf_idf, 0.9)) %>% #

ggplot(aes(reorder(word, tf_idf), tf_idf)) +

geom_col(fill = "skyblue") +

coord_flip() +

labs(x = "Word",

y = "TF-IDF",

title = "TF-IDF for Terms of Interest in Abstracts Related to Cefazolin and Penicillin Allergy")``` include a method like ``` mutate(TermOfInterest = ifelse(word %in% terms, "Yes", "No")) %>%``` and ``` geom_text(aes(label = ifelse(TermOfInterest == "Yes", as.character(word), "")), hjust = -0.1) +

scale_fill_manual(values = c("Yes" = "#fa7e1e", "No" = "#4f5bd5")) +`

**ChatGPT**

Sure, here is how you can create a TF-IDF plot highlighting the terms of interest. It uses the ```mutate()``` function to create a new column called "TermOfInterest" which indicates whether the word is in the terms list. The ```geom_text()``` function is used to add labels to the bars, and ```scale_fill_manual()``` is used to control the color of the bars based on whether the word is a term of interest.

```r

tf_idf %>%

filter(tf_idf > quantile(tf_idf, 0.9)) %>% # Optional: Only show words with highest TF-IDF values

mutate(TermOfInterest = ifelse(word %in% terms, "Yes", "No")) %>%

ggplot(aes(reorder(word, tf_idf), tf_idf, fill = TermOfInterest)) +

geom_col() +

coord_flip() +

geom_text(aes(label = ifelse(TermOfInterest == "Yes", as.character(word), "")), hjust = -0.1) +

scale_fill_manual(values = c("Yes" = "#fa7e1e", "No" = "#4f5bd5")) +

labs(x = "Word",

y = "TF-IDF",

title = "TF-IDF for Terms of Interest in Abstracts Related to Cefazolin and Penicillin Allergy",

fill = "Term of Interest")

```

In this plot, words in your term of interest will be highlighted with different colors and labels, while the remaining words will be displayed in a different color without labels.

**User**

I notice that our terms of interest appear in about 1% of all rows in tf_idf. I though that we would only have 1 row for every unique term

**ChatGPT**

In the TF-IDF calculation, a "document" usually refers to an individual text record - in this case, each individual abstract would typically be considered a document. The TF-IDF value is then calculated for each word in each document, which is why you may have multiple rows for the same word if it appears in multiple documents.

However, based on the code you've provided, it appears that all the abstracts have been combined into a single document. This may be why you are seeing only one row per term. If you wanted to treat each abstract as a separate document, you would need to adjust the data processing steps accordingly.

Here's how you might adjust the code to treat each abstract as a separate document:

```r

\# Split all_abstracts into a list of individual abstracts

individual_abstracts <- strsplit(all_abstracts, split = "\n\n")

\# Create a tibble with one row per abstract

abstracts_tibble <- tibble(abstract = unlist(individual_abstracts))

\# Add a document ID to each abstract

abstracts_tibble <- abstracts_tibble %>% mutate(document_id = row_number())

\# Tokenize the abstracts, which means to break the text into individual words:

tokenized_abstracts <- abstracts_tibble %>%

unnest_tokens(word, abstract)

\# Then you can proceed with the rest of the steps as before, but when calculating TF-IDF, use document_id as the document variable:

tf_idf <- tokenized_abstracts %>%

count(document_id, word) %>%

bind_tf_idf(word, document_id, n) %>%

arrange(desc(tf_idf))

```

Now, each row of the tf_idf data frame will represent a unique word-document combination, and you will have multiple rows for the same word if it appears in multiple documents.

**User**

can we remove the text labels on the y axis ticks

**ChatGPT**

Yes, it is possible to remove the text labels on the y-axis ticks by setting ```theme(axis.text.y=element_blank())``` in your ggplot. Here is an example with your code:

```r

tf_idf %>%

mutate(TermOfInterest = ifelse(word %in% terms, "Yes", "No")) %>%

filter(tf_idf > quantile(tf_idf, 0.9)) %>%

ggplot(aes(reorder(word, tf_idf), tf_idf, fill = TermOfInterest)) +

geom_col() +

coord_flip() +

geom_text(aes(label = ifelse(TermOfInterest == "Yes", as.character(word), "")), hjust = -0.1) +

scale_fill_manual(values = c("Yes" = "#fa7e1e", "No" = "#4f5bd5")) +

labs(x = "Word",

y = "TF-IDF",

title = "TF-IDF for Terms of Interest in Abstracts Related to Cefazolin and Penicillin Allergy") +

theme(axis.text.y=element_blank())

```

This will create a plot where the y-axis labels are removed while the other parts of the plot remain the same. The ```theme()``` function in ggplot2 allows you to adjust various elements of the plot's appearance, and ```element_blank()``` is used to turn off a particular element. In this case, we are turning off the y-axis text labels.

**User**

can we rank the y-axis based on ```tf_idf``` instead of ```word``` ```tf_idf %>%

arrange(desc(tf_idf)) %>%

\# filter(tf_idf > quantile(tf_idf, 0.9)) %>% # Optional: Only show words with highest TF-IDF values

mutate(TermOfInterest = ifelse(word %in% terms, "Yes", "No")) %>%

ggplot(aes(reorder(word, tf_idf), tf_idf, fill = TermOfInterest)) +

geom_col() +

coord_flip() +

\# geom_text(aes(label = ifelse(TermOfInterest == "Yes", as.character(word), "")), hjust = -0.1) +

scale_fill_manual(values = c("Yes" = "#fa7e1e", "No" = "#4f5bd5")) +

theme(axis.text.y=element_blank()) +

labs(x = "Word",

y = "TF-IDF",

title = "TF-IDF for Terms of Interest in Abstracts Related to Cefazolin and Penicillin Allergy",

fill = "Term of Interest")```

**ChatGPT**

Absolutely, you can rank the y-axis based on ```tf_idf``` instead of ```word``` by changing the ```reorder()``` function to reorder on ```tf_idf```:

Here's the adjusted code:

```r

tf_idf %>%

arrange(desc(tf_idf)) %>%

mutate(TermOfInterest = ifelse(word %in% terms, "Yes", "No")) %>%

ggplot(aes(reorder(word, -tf_idf), tf_idf, fill = TermOfInterest)) +

geom_col() +

coord_flip() +

scale_fill_manual(values = c("Yes" = "#fa7e1e", "No" = "#4f5bd5")) +

theme(axis.text.y=element_blank()) +

labs(x = "Word",

y = "TF-IDF",

title = "TF-IDF for Terms of Interest in Abstracts Related to Cefazolin and Penicillin Allergy",

fill = "Term of Interest")

```

In this case, the ```-tf_idf``` in ```reorder(word, -tf_idf)``` is used to sort the terms in descending order of their tf-idf values, since ```reorder()``` sorts in ascending order by default. As a result, the terms with the highest tf-idf values will be at the top of the y-axis.

**User**

i still appears as though the terms are grouped such that some values of high and low value are next to each other instead of complete order

**ChatGPT**

My apologies for the confusion earlier. When dealing with factors, the ```reorder()``` function rearranges the levels of the factor in ascending order based on the second argument provided to it. When using ```coord_flip()```, the plot will start from the bottom and move up. So in this case, lower tf-idf values are plotted first and higher ones are plotted last, and the plot is flipped horizontally.

But as we want to sort tf-idf values in descending order, we should reverse the factor levels manually after they've been reordered. Let's adjust the code accordingly:

```r

tf_idf %>%

arrange(desc(tf_idf)) %>%

mutate(TermOfInterest = ifelse(word %in% terms, "Yes", "No")) %>%

mutate(word = factor(word, levels = rev(unique(word)))) %>% # Reverse the factor levels

ggplot(aes(x = word, y = tf_idf, fill = TermOfInterest)) +

geom_col() +

coord_flip() +

scale_fill_manual(values = c("Yes" = "#fa7e1e", "No" = "#4f5bd5")) +

theme(axis.text.y=element_blank()) +

labs(x = "Word",

y = "TF-IDF",

title = "TF-IDF for Terms of Interest in Abstracts Related to Cefazolin and Penicillin Allergy",

fill = "Term of Interest")

```

This should ensure that the bars are plotted in descending order of tf-idf values from top to bottom.

**User**

can we make a new column of numbers based on the oder if we rank the data based on values in column tf_idf

**ChatGPT**

Absolutely. The ```row_number()``` function in dplyr can be used for this purpose. It assigns a unique row number to each row of a data frame, based on the order of the rows. If you use it after ```arrange(desc(tf_idf))```, it will assign row numbers based on the descending order of ```tf_idf``` values.

Here's how to do it:

```r

tf_idf %>%

arrange(desc(tf_idf)) %>%

mutate(TermOfInterest = ifelse(word %in% terms, "Yes", "No")) %>%

mutate(Rank = row_number()) %>% # Add a new 'Rank' column

ggplot(aes(x = Rank, y = tf_idf, fill = TermOfInterest)) +

geom_col() +

scale_fill_manual(values = c("Yes" = "#fa7e1e", "No" = "#4f5bd5")) +

labs(x = "Rank",

y = "TF-IDF",

title = "TF-IDF for Terms of Interest in Abstracts Related to Cefazolin and Penicillin Allergy",

fill = "Term of Interest")

```

In this plot, the x-axis represents the rank of each word, based on ```tf_idf```. The bars are plotted from left to right in order of descending ```tf_idf``` values. The color of each bar indicates whether the word is a term of interest.

**User**

can we add ``` geom_text(aes(label = ifelse(TermOfInterest == "Yes", as.character(word), "")), hjust = -0.1) +``` but rotate it 90 degress

**ChatGPT**

Yes, the ```geom_text()``` function in ggplot2 allows for text rotation. Here's how to do it:

```r

tf_idf %>%

arrange(desc(tf_idf)) %>%

mutate(TermOfInterest = ifelse(word %in% terms, "Yes", "No")) %>%

mutate(Rank = row_number()) %>% # Add a new 'Rank' column

ggplot(aes(x = Rank, y = tf_idf, fill = TermOfInterest)) +

geom_col() +

geom_text(aes(label = ifelse(TermOfInterest == "Yes", as.character(word), "")), angle = 90, hjust = -0.1) +

scale_fill_manual(values = c("Yes" = "#fa7e1e", "No" = "#4f5bd5")) +

labs(x = "Rank",

y = "TF-IDF",

title = "TF-IDF for Terms of Interest in Abstracts Related to Cefazolin and Penicillin Allergy",

fill = "Term of Interest")

```

The ```angle``` argument inside ```geom_text()``` is used to rotate the labels. An angle of 90 degrees rotates the labels 90 degrees counter-clockwise.

**User**

can we use geom repel text to make these more readable ``` geom_text(aes(label = ifelse(TermOfInterest == "Yes", as.character(word), "")), angle = 90, hjust = -0.1, size =1.5) +`

**ChatGPT**

Sure, you can replace ```geom_text``` with ```geom_text_repel``` from the ```ggrepel``` package to automatically prevent the text labels from overlapping. It's a great tool when dealing with a cluttered plot.

If you have not installed the package, do so by running ```install.packages("ggrepel")```.

Here is the updated code:

```r

\# Make sure to load the library

library(ggrepel)

tf_idf %>%

arrange(desc(tf_idf)) %>%

mutate(TermOfInterest = ifelse(word %in% terms, "Yes", "No")) %>%

mutate(Rank = row_number()) %>% # Add a new 'Rank' column

ggplot(aes(x = Rank, y = tf_idf, fill = TermOfInterest)) +

geom_col() +

geom_text_repel(aes(label = ifelse(TermOfInterest == "Yes", as.character(word), "")),

angle = 90, hjust = -0.1, size =1.5) +

scale_fill_manual(values = c("Yes" = "#fa7e1e", "No" = "#4f5bd5")) +

labs(x = "Rank",

y = "TF-IDF",

title = "TF-IDF for Terms of Interest in Abstracts Related to Cefazolin and Penicillin Allergy",

fill = "Term of Interest")

```

The ```geom_text_repel()``` function will automatically try to position labels in a way that avoids overlap. Adjusting the parameters of ```geom_text_repel()``` might be necessary to get a pleasing result, as the effectiveness can depend on the specifics of your plot.

**User**

Great, now lets do a correlation Plot or Network Analysis - here is the current code block: ```# Start time: 2023-07-31 12:50

\# Pubmed knowledge ----

library(rentrez)

library(XML)

\# Define your search terms

terms <- c("cefazolin", "penicillin", "allergy")

\# Create the query string by joining the terms with '[Abstract] AND '

query <- paste0(terms, "[Abstract]", collapse = " AND ")

\# Search for publications in PubMed

papers <- entrez_search(db="pubmed", term=query)

\# Fetch records in "abstract" format

xmlData <- entrez_fetch(db="pubmed", id=papers$ids, rettype="abstract", retmode="xml")

\# Parse XML

doc <- xmlParse(xmlData)

\# Extract abstract nodes

abstractNodes <- getNodeSet(doc, "//AbstractText")

\# Extract the content of abstract nodes

abstracts <- sapply(abstractNodes, xmlValue)

\# Combine all abstracts into a single string, separated by two newline characters

all_abstracts <- paste(abstracts, collapse = "\n\n")

\# Write all abstracts to a single file

writeLines(all_abstracts, "../data/all_abstracts.txt")

\# Plot term frequency data ----

#install.packages(c("tidytext", "ggplot2", "stopwords"))

library(tidyverse)

library(tidytext)

library(ggplot2)

library(ggrepel)

library(stopwords)

\# Create a tibble

abstracts_tibble <- tibble(abstract = all_abstracts)

\# Tokenize the abstracts, which means to break the text into individual words:

tokenized_abstracts <- abstracts_tibble %>%

unnest_tokens(word, abstract)

\# Then you can proceed with the rest of the steps as before.

\# Remove common "stop words" (like "and", "the", "of", etc.) as they are usually not informative:

data(stop_words)

tokenized_abstracts <- tokenized_abstracts %>%

anti_join(stop_words)

\# Count the frequency of each word:

word_counts <- tokenized_abstracts %>%

count(word, sort = TRUE)

\# Make the plot, showing the most common words:

word_counts %>%

filter(n > quantile(n, 0.9)) %>% # Optional: Only show the most common words

ggplot(aes(reorder(word, n), n)) +

geom_col() +

coord_flip() +

labs(x = "Word",

y = "Frequency",

title = "Most Common Words in Abstracts Related to Cefazolin and Penicillin Allergy")

\# Define the terms of interest

terms <- c("cefazolin", "penicillin", "allergy")

\# Make the plot, showing the most common words:

plot <- word_counts %>%

mutate(TermOfInterest = ifelse(word %in% terms, "Yes", "No")) %>%

filter(n > quantile(n, 0.9)) %>% # Optional: Only show the most common words

ggplot(aes(reorder(word, n), n, fill = TermOfInterest)) +

geom_col() +

geom_text(aes(label = ifelse(TermOfInterest == "Yes", as.character(word), "")), hjust = -0.1) +

scale_fill_manual(values = c("Yes" = "#fa7e1e", "No" = "#4f5bd5")) +

coord_flip() +

theme_bw() +

labs(x = "Word",

y = "Frequency",

title = "Most common words in \nabstracts related to cefazolin and penicillin allergy",

fill = "Term of interest")

\# Save it as a PDF

ggsave(filename = "../data/abstracts_word_frequency.pdf", plot = plot, width = 8, height = 12, units = "in")

\# Plot TF-IDF ----

\# Split the single string of all_abstracts into individual abstracts

abstracts_tibble <- tibble(abstract = strsplit(all_abstracts, "\n\n")[[1]])

\# Add a unique identifier for each abstract:

abstracts_tibble <- abstracts_tibble %>%

mutate(abstract_id = row_number())

\# Tokenize the abstracts with the unique identifier:

tokenized_abstracts <- abstracts_tibble %>%

unnest_tokens(word, abstract)

\# Remove common "stop words" (like "and", "the", "of", etc.) as they are usually not informative:

data(stop_words)

tokenized_abstracts <- tokenized_abstracts %>%

anti_join(stop_words)

\# Calculate TF-IDF:

tf_idf <- tokenized_abstracts %>%

count(abstract_id, word) %>%

bind_tf_idf(word, abstract_id, n) %>%

arrange(desc(tf_idf))

\# plot

plot2 <- tf_idf %>%

arrange(desc(tf_idf)) %>%

mutate(TermOfInterest = ifelse(word %in% terms, "Yes", "No")) %>%

mutate(Rank = row_number()) %>% # Add a new 'Rank' column

ggplot(aes(x = Rank, y = tf_idf, fill = TermOfInterest)) +

geom_col() +

scale_fill_manual(values = c("Yes" = "#fa7e1e", "No" = "#4f5bd5")) +

geom_text_repel(aes(label = ifelse(TermOfInterest == "Yes", as.character(word), "")),

max.overlaps = 200,

alpha = .8,

angle = 90, hjust = -1.2, size =2) +

theme_bw() +

labs(x = "Rank",

y = "TF-IDF",

title = "TF-IDF for terms of interest in \nabstracts related to cefazolin and penicillin allergy",

fill = "Term of interest")

\# Save it as a PDF

ggsave(filename = "../data/abstracts_tf_idf.pdf", plot = plot2, width = 10, height = 8, units = "in")

```

**ChatGPT**

For a correlation plot or network analysis, you will need to compute a correlation matrix where each cell represents a measure of the association between two terms. The first step is to construct a document-term matrix (DTM) or a term co-occurrence matrix.

For this, we will use the ```tm``` and ```textTinyR``` libraries. The ```tm``` library provides the function ```DocumentTermMatrix()``` which generates a DTM and ```textTinyR``` allows us to compute a term co-occurrence matrix.

Please note that these are resource-intensive processes and may take some time for large corpora. Here is a basic example of how to construct such a plot:

First, we need to install the necessary packages if you haven't done so:

```r

install.packages(c("tm", "textTinyR", "reshape2", "corrr", "igraph", "ggraph"))

```

Then, load the required libraries:

```r

library(tm)

library(textTinyR)

library(reshape2)

library(corrr)

library(igraph)

library(ggraph)

```

Then, generate a Document Term Matrix (DTM) using the ```tm``` library:

```r

\# Generate a corpus from the abstracts vector

corpus <- Corpus(VectorSource(abstracts))

\# Create the Document-Term Matrix

dtm <- DocumentTermMatrix(corpus)

\# Convert the DTM to a regular matrix

dtm_mat <- as.matrix(dtm)

\# Compute term correlations

term_cor <- cor(dtm_mat)

```

We then transform the correlation matrix into a graph object that we can use to generate the network plot:

```r

\# Find the correlations that are above a certain threshold (e.g., 0.7)

term_cor_melted <- melt(cor(term_cor)) %>%

filter(value > 0.7)

\# Create a graph object

graph <- graph_from_data_frame(term_cor_melted)

\# Generate the network plot

ggraph(graph, layout = 'fr') +

geom_edge_link(aes(edge_alpha = value), show.legend = FALSE) +

geom_node_point(size = 5) +

geom_node_text(aes(label = name), vjust = 1, hjust = 1) +

theme_void()

```

You may need to adjust the parameters and thresholds to get a graph that makes sense for your specific data.

**User**

I cannot install library(tm). lets do it another way

**ChatGPT**

Understood. If you can't install the ```tm``` package, another way to do this is using the ```tidytext``` package to create a co-occurrence matrix, then use this matrix to calculate the correlations, and finally use ```igraph``` and ```ggraph``` for plotting.

Let's proceed as follows:

First, let's compute a co-occurrence matrix. In this case, we are considering two words co-occur if they appear in the same abstract:

```r

library(dplyr)

library(tidyr)

library(tidytext)

\# Compute a co-occurrence matrix

co_occur <- tokenized_abstracts %>%

pairwise_count(word, abstract_id, sort = TRUE, upper = FALSE)

```

Next, we need to convert this to a format suitable for generating a correlation network plot. We can use the ```igraph``` package for this:

```r

library(igraph)

\# Create an igraph object

g <- graph_from_data_frame(co_occur)

\# Calculate the edge weights based on the frequency of co-occurrence

E(g)$weight <- E(g)$n

\# Prune the graph to only keep edges with weights above a certain threshold

g <- delete_edges(g, E(g)[E(g)$weight < quantile(E(g)$weight, 0.95)])

\# Calculate the layout of the graph

layout <- layout_with_fr(g)

```

Finally, we can create a network plot using the ```ggraph``` package:

```r

library(ggraph)

\# Generate the network plot

ggraph(g, layout = layout) +

geom_edge_link(aes(edge_alpha = weight), edge_width = 0.1, show.legend = FALSE) +

geom_node_point(size = 3) +

geom_node_text(aes(label = name), repel = TRUE) +

theme_void()

```

Please note that you may need to adjust the threshold in the ```delete_edges``` function depending on the size of your dataset and the level of detail you want in your network plot.

**User**

> # Compute a co-occurrence matrix

> co_occur <- tokenized_abstracts %>%

+	pairwise_count(word, abstract_id, sort = TRUE, upper = FALSE)

Error in pairwise_count(., word, abstract_id, sort = TRUE, upper = FALSE) :

could not find function "pairwise_count"

**ChatGPT**

I apologize for the confusion, the function ```pairwise_count()``` is actually a part of the ```widyr``` package in R, which provides functions to work with wide-format data in a tidy way.

Here is how you can correct the error:

```R

install.packages("widyr")

library(widyr)

\# Compute a co-occurrence matrix

co_occur <- tokenized_abstracts %>%

pairwise_count(word, abstract_id, sort = TRUE, upper = FALSE)

```

Then proceed with the rest of the code as mentioned earlier. The ```widyr``` package is designed to work well with ```tidytext``` and ```dplyr``` packages, which you're already using in your script.

**User**

is there other plots that can demonstrate the data in ggplot

**ChatGPT**

There are several types of plots that you can make with ggplot2. Here are a few examples that might be relevant for your dataset:

1. **Correlation plot:** This plot is a great way to visualize correlations between different variables. For example, if you want to see the correlation between the frequency of different terms in your abstracts, you can calculate a correlation matrix and use ggplot2 to visualize it.

2. **Heatmap:** A heatmap can be used to visualize a matrix of data (like a co-occurrence matrix) by color-coding the cells according to their values. This might be a good way to visualize your co-occurrence data.

3. **Network plot:** A network plot visualizes relationships between different entities (like words). You can create a network plot using the ggraph package, which is built on top of ggplot2.

For instance, if you want to create a heatmap using your co-occurrence matrix, you could use the following code:

```r

library(ggplot2)

library(reshape2)

\# First, we need to reshape the data to a long format

co_occur_long <- melt(co_occur)

ggplot(co_occur_long, aes(x = Var1, y = Var2, fill = value)) +

geom_tile() +

theme(axis.text.x = element_text(angle = 90, hjust = 1)) +

labs(x = "Word 1",

y = "Word 2",

fill = "Co-occurrence",

title = "Heatmap of Term Co-occurrence in Abstracts Related to Cefazolin and Penicillin Allergy")

```

Please remember that you might need to preprocess your data to get the required format for these plots.

**User**

we have a lot of datapoints. is it possible to remove very low information content? ```co_occur_long |>

head(200) |>

ggplot(aes(x = item1, y = item2, fill = value)) +

geom_tile() +

theme(axis.text.x = element_text(angle = 90, hjust = 1)) +

labs(x = "Word 1",

y = "Word 2",

fill = "Co-occurrence",

title = "Heatmap of Term Co-occurrence in Abstracts Related to Cefazolin and Penicillin Allergy")

```

**ChatGPT**

If you're dealing with a large amount of data and want to remove low information content, you could apply a filter to only include word pairs that co-occur above a certain threshold. Here's how you can do this:

```r

\# Filter to include only word pairs that co-occur more than a certain number of times

threshold <- 10 # Change this to suit your needs

co_occur_long <- co_occur_long %>% filter(value > threshold)

\# Plot

ggplot(co_occur_long, aes(x = item1, y = item2, fill = value)) +

geom_tile() +

theme(axis.text.x = element_text(angle = 90, hjust = 1)) +

labs(x = "Word 1",

y = "Word 2",

fill = "Co-occurrence",

title = "Heatmap of Term Co-occurrence in Abstracts Related to Cefazolin and Penicillin Allergy")

```

In this code, ```threshold``` is the minimum number of co-occurrences that a word pair needs to be included in the plot. You can adjust this to suit your needs.

Keep in mind that depending on your data, setting the threshold too high might exclude many word pairs, while setting it too low might include too much noise in the plot. You might need to experiment with different values to find the one that works best for your data.

**User**

lets make the x-axis labels rotate 45 ```# Plot

ggplot(co_occur_long, aes(x = item1, y = item2, fill = value)) +

geom_tile() +

theme(axis.text.x = element_text(angle = 90, hjust = 1)) +

theme_bw() +

labs(x = "Word 1",

y = "Word 2",

fill = "Co-occurrence",

title = "Heatmap of term co-occurrence in \nabstracts related to cefazolin and penicillin allergy")`

**ChatGPT**

Absolutely, you can modify the ```element_text()``` argument to change the angle to 45 degrees. Here's how:

```r

\# Plot

ggplot(co_occur_long, aes(x = item1, y = item2, fill = value)) +

geom_tile() +

theme(axis.text.x = element_text(angle = 45, hjust = 1)) +

theme_bw() +

labs(x = "Word 1",

y = "Word 2",

fill = "Co-occurrence",

title = "Heatmap of term co-occurrence in \nabstracts related to cefazolin and penicillin allergy")

```

This will make the x-axis labels rotate 45 degrees.

**User**

in this plot, we have too many labels. could we just add the ones which have shortest edges ```# Generate the network plot

ggraph(g, layout = layout) +

geom_edge_link(aes(edge_alpha = weight), edge_width = 0.1, show.legend = FALSE) +

geom_node_point(size = 1) +

geom_node_text(aes(label = name), repel = TRUE) +

theme_void()`

**ChatGPT**

To limit the labels in the network graph to only the nodes with the highest degrees (i.e., connected to most other nodes), you'll need to adjust the ```geom_node_text()``` function to only include those nodes.

Here's how you can modify your code:

```r

\# Calculate degrees of each node

degrees <- igraph::degree(g)

\# Plot

ggraph(g, layout = layout) +

geom_edge_link(aes(edge_alpha = weight), edge_width = 0.1, show.legend = FALSE) +

geom_node_point(size = 1) +

geom_node_text(data = degrees[degrees > quantile(degrees, 0.9)], aes(label = name), repel = TRUE) +

theme_void()

```

In this case, ```quantile(degrees, 0.9)``` selects the top 10% of nodes based on their degrees. You can adjust this quantile to suit your needs.

However, keep in mind that ```geom_node_text()``` needs a dataframe as input. Thus, you may have to create a new dataframe that only includes the nodes you're interested in.

Let's try to do this:

```r

\# Create a dataframe of nodes with their corresponding degrees

df_degrees <- data.frame(name = names(degrees), degree = degrees)

\# Plot

ggraph(g, layout = layout) +

geom_edge_link(aes(edge_alpha = weight), edge_width = 0.1, show.legend = FALSE) +

geom_node_point(size = 1) +

geom_node_text(data = df_degrees %>%

filter(degree > quantile(degree, 0.9)),

aes(label = name), repel = TRUE) +

theme_void()

```

This version of the code should limit the labels to only the nodes with the highest degrees.

**User**

> # Plot

> ggraph(g, layout = layout) +

+	geom_edge_link(aes(edge_alpha = weight), edge_width = 0.1, show.legend = FALSE) +

+	geom_node_point(size = 1) +

+	geom_node_text(data = degrees[degrees > quantile(degrees, 0.9)], aes(label = name), repel = TRUE) +

+	theme_void()

Error in ```fortify()```:

! ```data``` must be a <data.frame>, or an

object coercible by ```fortify()```, not a double

vector.

Run ```rlang::last_trace()``` to see where the error occurred.

> # Create a dataframe of nodes with their corresponding degrees

> df_degrees <- data.frame(name = names(degrees), degree = degrees)

> # Plot

> ggraph(g, layout = layout) +

+	geom_edge_link(aes(edge_alpha = weight), edge_width = 0.1, show.legend = FALSE) +

+	geom_node_point(size = 1) +

+	geom_node_text(data = df_degrees %>%

+	filter(degree > quantile(degree, 0.9)),

+	aes(label = name), repel = TRUE) +

+	theme_void()

Error in ```geom_node_text()```:

! Problem while computing aesthetics.

ℹ Error occurred in the 3rd layer.

Caused by error in ```FUN()```:

! object 'x' not found

**ChatGPT**

Apologies for the confusion. It seems I made an error. The ```geom_node_text()``` function doesn't accept a separate dataframe for the ```data``` argument. Let's approach this differently. We'll mark nodes of interest with a special attribute, and then use this attribute to conditionally display labels.

```r

\# Add a node attribute based on degree

V(g)$high_degree <- degree(g) > quantile(degree(g), 0.9)

\# Plot

ggraph(g, layout = layout) +

geom_edge_link(aes(edge_alpha = weight), edge_width = 0.1, show.legend = FALSE) +

geom_node_point(size = 1) +

geom_node_text(aes(label = ifelse(high_degree, name, "")), repel = TRUE) +

theme_void()

```

This code first adds an attribute to each node in the graph indicating whether it's a high degree node or not. Then, in the ```geom_node_text()``` function, it conditionally displays labels based on this attribute.

Again, ```quantile(degree(g), 0.9)``` is used to select the top 10% of nodes by degree, and you can adjust this quantile to suit your needs.

**User**

can i save this ```ggraph``` to pdf

**ChatGPT**

Yes, you can save your ```ggraph``` plot to a PDF using the ```ggsave()``` function just like you would do for a ```ggplot2``` graph. Here is an example:

```r

\# Create the plot

plot <- ggraph(g, layout = layout) +

geom_edge_link(aes(edge_alpha = weight), edge_width = 0.1, show.legend = FALSE) +

geom_node_point(size = 1) +

geom_node_text(aes(label = ifelse(high_degree, name, "")), repel = TRUE) +

theme_void()

\# Save the plot

ggsave(filename = "../data/network_plot.pdf", plot = plot, width = 10, height = 10, units = "in")

```

In the above code, replace ```"../data/network_plot.pdf"``` with your preferred file path and name. The ```width``` and ```height``` parameters specify the dimensions of the output PDF, and you can adjust these values as necessary. The ```units``` parameter can be ```"in"``` for inches, ```"cm"``` for centimeters, or ```"mm"``` for millimeters.

**User**

this iso good but we have one cluster of nodes that are highly related, but we cannot see any labels. how can we check what value to use to determine text labellening ```# Plot

plot4 <- ggraph(g, layout = layout) +

geom_edge_link(aes(edge_alpha = weight),

edge_width = 0.5, show.legend = FALSE) +

geom_node_point(size = .3, alpha = 0.5) +

geom_node_text(aes(label = ifelse(high_degree, name, "")), repel = TRUE) +

theme_void()`

**ChatGPT**

If you have one cluster of nodes that are highly related and the text labels are overlapping or not visible, one approach could be to adjust the threshold for ```high_degree```. The threshold that I used was the 90th percentile (```quantile(degrees, 0.9)```), but you can adjust this value to be higher or lower as needed. Here is an example:

```r

\# Adjust the high_degree threshold

high_degree <- degrees > quantile(degrees, 0.95) # Change to the 95th percentile

\# Plot

plot <- ggraph(g, layout = layout) +

geom_edge_link(aes(edge_alpha = weight), edge_width = 0.5, show.legend = FALSE) +

geom_node_point(size = 0.3, alpha = 0.5) +

geom_node_text(aes(label = ifelse(high_degree, name, "")), repel = TRUE) +

theme_void()

\# Save the plot

ggsave(filename = "../data/network_plot.pdf", plot = plot, width = 10, height = 10, units = "in")

```

By increasing the percentile, fewer nodes will be considered as high degree and therefore fewer labels will be displayed. You might need to try a few different values to find the one that works best for your specific network. Another option is to use a different method to identify the nodes of interest, depending on the specific characteristics of your network and the research questions you are interested in. For example, you could label the nodes that are part of the largest connected component, or the nodes that have the highest betweenness centrality.

**User**

in this plot what would the X and Y axis values be characterised as?

**ChatGPT**

In the network plot generated using ```ggraph```, the X and Y axis values represent the positions of the nodes in the two-dimensional layout of the network. The layout algorithm determines the positions of the nodes in a way that tries to visually represent the relationships and connections between nodes in the network.

In the code provided, the ```layout``` variable is used to specify the layout algorithm. The layout algorithm calculates the two-dimensional positions of the nodes based on the structure of the network and the relationships between nodes. Some common layout algorithms used in network visualization include the Fruchterman-Reingold layout, the Kamada-Kawai layout, and the circular layout.

The X and Y coordinates of the nodes represent the positions of the nodes in the layout. Nodes that are close to each other in the layout are likely to be more connected in the network, while nodes that are far apart are less connected. The layout algorithm aims to optimize the arrangement of nodes to make the network structure more visually informative and easier to interpret.

Keep in mind that the X and Y axis values themselves do not have specific meaning beyond representing the positions of the nodes in the layout. The real information lies in the connections between nodes, which are represented by the edges in the network plot. The layout algorithm aims to arrange the nodes in a way that makes it easier to visually identify patterns and structures in the network, such as clusters, hubs, and subgroups.

**User**

how do we count the number of abstracts used here: ```# Start time: 2023-07-31 12:50

\# End time: 2023-07-31 14:43

\# Pubmed knowledge ----

library(rentrez)

library(XML)

\# Define your search terms

terms <- c("cefazolin", "penicillin", "allergy")

\# Create the query string by joining the terms with '[Abstract] AND '

query <- paste0(terms, "[Abstract]", collapse = " AND ")

\# Search for publications in PubMed

papers <- entrez_search(db="pubmed", term=query)

\# Fetch records in "abstract" format

xmlData <- entrez_fetch(db="pubmed", id=papers$ids, rettype="abstract", retmode="xml")

\# Parse XML

doc <- xmlParse(xmlData)

\# Extract abstract nodes

abstractNodes <- getNodeSet(doc, "//AbstractText")

\# Extract the content of abstract nodes

abstracts <- sapply(abstractNodes, xmlValue)

\# Combine all abstracts into a single string, separated by two newline characters

all_abstracts <- paste(abstracts, collapse = "\n\n")

```

**ChatGPT**

To count the number of abstracts used, you can simply check the length of the ```papers$ids``` vector, which contains the unique identifiers (PMIDs) of the retrieved publications from PubMed. Each PMID corresponds to an individual abstract that matches the search terms.

Here's how you can count the number of abstracts used in the provided code:

```R

\# Search for publications in PubMed

papers <- entrez_search(db = "pubmed", term = query)

\# Get the number of abstracts used

num_abstracts <- length(papers$ids)

\# Print the number of abstracts

print(num_abstracts)

```

After running this code, you will see the total number of abstracts used in the search printed in the console.

**User**

since pubmed IDs, such as "PMID: 37363051", can be searched for by appending the number to the main URL like so "https://pubmed.ncbi.nlm.nih.gov/37363051/" we would automatically print all of the URLs for the PMID held in ```papers$ids``` by appending each to the URL ```https://pubmed.ncbi.nlm.nih.gov/`

**ChatGPT**

You can use the ```paste``` function in R to concatenate the main URL "https://pubmed.ncbi.nlm.nih.gov/" with each PMID in the ```papers$ids``` vector. This will create the full URLs for each PMID.

Here's how you can print all the URLs for the PMIDs:

```R

\# Concatenate the main URL with each PMID

pmid_urls <- paste0("https://pubmed.ncbi.nlm.nih.gov/", papers$ids)

\# Print all the URLs

print(pmid_urls)

```

After running this code, you will see a list of URLs, each corresponding to a specific PMID from the ```papers$ids``` vector. You can click on these URLs to access the corresponding abstracts on the PubMed website.

**User**

can i print those without list numbers and quotes

**ChatGPT**

Yes, you can use the ```cat``` function in R to print the URLs without list numbers and quotes. The ```cat``` function concatenates and prints the URLs as a continuous output.

Here's how you can print the URLs without list numbers and quotes:

```R

\# Concatenate the main URL with each PMID

pmid_urls <- paste0("https://pubmed.ncbi.nlm.nih.gov/", papers$ids)

\# Print all the URLs without list numbers and quotes

cat(pmid_urls, sep = "\n")

```

The ```sep = "\n"``` argument in the ```cat``` function specifies that each URL should be separated by a newline character, so they will be printed on separate lines. This will give you a clean output with one URL per line.
