## Supplemental for "Reply to Dages, et al. You AIn’t using it right: Artificial intelligence progress in allergy": jaci_gpt.R.txt

# Start time: 2023-07-31 12:50
# End time:  2023-07-31 14:43

# Pubmed knowledge ----
library(rentrez)
library(XML)

# Define your search terms
terms <- c("cefazolin", "penicillin", "allergy")

# Create the query string by joining the terms with '[Abstract] AND '
query <- paste0(terms, "[Abstract]", collapse = " AND ")

# Search for publications in PubMed
papers <- entrez_search(db = "pubmed", term = query)

# Fetch records in "abstract" format
xmlData <-
  entrez_fetch(
    db = "pubmed",
    id = papers$ids,
    rettype = "abstract",
    retmode = "xml"
  )

# Parse XML
doc <- xmlParse(xmlData)

# Extract abstract nodes
abstractNodes <- getNodeSet(doc, "//AbstractText")

# Extract the content of abstract nodes
abstracts <- sapply(abstractNodes, xmlValue)

# Combine all abstracts into a single string, separated by two newline characters
all_abstracts <- paste(abstracts, collapse = "\n\n")

# Write all abstracts to a single file
writeLines(all_abstracts, "../data/all_abstracts.txt")

# Plot term frequency data ----
library(tidytext)
library(ggplot2)
library(ggrepel)
library(stopwords)
library(tidyverse)

# Create a tibble
abstracts_tibble <- tibble(abstract = all_abstracts)

# Tokenize the abstracts, which means to break the text into individual words:
tokenized_abstracts <- abstracts_tibble %>%
  unnest_tokens(word, abstract)

# Remove common "stop words" (like "and", "the", "of", etc.) as they are usually not informative:
data(stop_words)
tokenized_abstracts <- tokenized_abstracts %>%
  anti_join(stop_words)

# Count the frequency of each word:
word_counts <- tokenized_abstracts %>%
  count(word, sort = TRUE)

# Make the plot, showing the most common words:
word_counts %>%
  filter(n > quantile(n, 0.9)) %>%  # Optional: Only show the most common words
  ggplot(aes(reorder(word, n), n)) +
  geom_col() +
  coord_flip() +
  labs(x = "Word",
       y = "Frequency",
       title = "Most Common Words in Abstracts Related to Cefazolin and Penicillin Allergy")

# Define the terms of interest
terms <- c("cefazolin", "penicillin", "allergy")

# Make the plot, showing the most common words:
title1 <-
  "Most common words in \nabstracts related to cefazolin and penicillin allergy (quantile 0.95)"
plot1 <- word_counts %>%
  mutate(TermOfInterest = ifelse(word %in% terms, "Yes", "No")) %>%
  filter(n > quantile(n, 0.95)) %>%  # Optional: Only show the most common words
  ggplot(aes(reorder(word, n), n, fill = TermOfInterest)) +
  geom_col() +
  geom_text(aes(label = ifelse(
    TermOfInterest == "Yes", as.character(word), ""
  )), hjust = -0.1) +
  scale_fill_manual(values = c("Yes" = "#fa7e1e", "No" = "#4f5bd5")) +
  coord_flip() +
  theme_bw() +
  labs(x = "Word",
       y = "Frequency",
       # title = title1,
       fill = "Term of interest")

# Save it as a PDF
ggsave(
  filename = "../data/abstracts_word_frequency.pdf",
  plot = plot1,
  width = 8,
  height = 12,
  units = "in"
)


# Plot TF-IDF ----
# Split the single string of all_abstracts into individual abstracts
abstracts_tibble <-
  tibble(abstract = strsplit(all_abstracts, "\n\n")[[1]])

# Add a unique identifier for each abstract:
abstracts_tibble <- abstracts_tibble %>%
  mutate(abstract_id = row_number())

# Tokenize the abstracts with the unique identifier:
tokenized_abstracts <- abstracts_tibble %>%
  unnest_tokens(word, abstract)

# Remove common "stop words" (like "and", "the", "of", etc.) as they are usually not informative:
data(stop_words)
tokenized_abstracts <- tokenized_abstracts %>%
  anti_join(stop_words)

# Calculate TF-IDF:
tf_idf <- tokenized_abstracts %>%
  count(abstract_id, word) %>%
  bind_tf_idf(word, abstract_id, n) %>%
  arrange(desc(tf_idf))

# plot
title2 <-
  "TF-IDF for terms of interest in \nabstracts related to cefazolin and penicillin allergy"

plot2 <- tf_idf %>%
  arrange(desc(tf_idf)) %>%
  mutate(TermOfInterest = ifelse(word %in% terms, "Yes", "No")) %>%
  mutate(Rank = row_number()) %>%  # Add a new 'Rank' column
  ggplot(aes(x = Rank, y = tf_idf, fill = TermOfInterest)) +
  geom_col() +
  scale_fill_manual(values = c("Yes" = "#fa7e1e", "No" = "#4f5bd5")) +
  geom_text_repel(
    aes(label = ifelse(
      TermOfInterest == "Yes", as.character(word), ""
    )),
    max.overlaps = 10,
    alpha = .8,
    angle = 90,
    hjust = -40,
    vjust = 4,
    size = 4
  ) +
  theme_bw() +
  labs(x = "Rank",
       y = "TF-IDF",
       # title = title2,
       fill = "Term of interest")

# Save it  as a PDF
ggsave(
  filename = "../data/abstracts_tf_idf.pdf",
  plot = plot2,
  width = 10,
  height = 8,
  units = "in"
)


# Plot correlation ----
library(dplyr)
library(tidyr)
# library(tidytext)
library(widyr)
library(igraph)
library(ggraph)

# Compute a co-occurrence matrix
co_occur <- tokenized_abstracts %>%
  pairwise_count(word, abstract_id, sort = TRUE, upper = FALSE)

# Create an igraph object
g <- graph_from_data_frame(co_occur)

# Calculate the edge weights based on the frequency of co-occurrence
E(g)$weight <- E(g)$n

# Prune the graph to only keep edges with weights above a certain threshold
g <-
  delete_edges(g, E(g)[E(g)$weight < quantile(E(g)$weight, 0.95)])

# Calculate the layout of the graph
layout <- layout_with_fr(g)

# Add a node attribute based on degree
V(g)$high_degree <- degree(g) > quantile(degree(g), 0.98)

# Plot
title3 <-
  "Network plot of edge weights based \non the frequency of co-occurrence in\nabstracts related to cefazolin and penicillin allergy (quantile  0.98)"
plot3 <- ggraph(g, layout = layout) +
  geom_edge_link(aes(edge_alpha = weight),
                 edge_width = 0.5,
                 show.legend = FALSE) +
  geom_node_point(size = .3, alpha = 0.5) +
  geom_node_text(
    aes(label = ifelse(high_degree, name, "")),
    color = "#4f5bd5",
    repel = TRUE,
    max.overlaps = 50,
    hjust = -0.6,
    size = 4
  ) +
  theme_void()
# labs(title = title3, size=0.6)

# Save it  as a PDF
ggsave(
  filename = "../data/network_plot.pdf",
  plot = plot3,
  width = 6,
  height = 6,
  units = "in"
)

# Plot heatmap ----
library(ggplot2)
library(reshape2)

# First, we need to reshape the data to a long format
co_occur_long <- melt(co_occur)

# Filter to include only word pairs that co-occur more than a certain number of times
threshold <- 9  # Change this to suit your needs
co_occur_long <- co_occur_long %>% filter(value > threshold)

# Plot
title4 <-
  "Heatmap of term co-occurrence in \nabstracts related to cefazolin and penicillin allergy (co-occurrence threshold >9)"

plot4 <-
  ggplot(co_occur_long, aes(x = item1, y = item2, fill = value)) +
  geom_tile() +
  theme_bw() +
  theme(axis.text.x = element_text(angle = 45, hjust = 1)) +
  labs(
    x = "Word 1",
    y = "Word 2",
    fill = "Co-occurrence")
# title = title4

# Save it  as a PDF
ggsave(
  filename = "../data/abstracts_heatmap.pdf",
  plot = plot4,
  width = 10,
  height = 8,
  units = "in"
)

# patchwork ----
library(patchwork)
patch0 <- (plot1 | plot2) / (plot3 | plot4) + plot_annotation(tag_levels = 'A')

patch1 <- (plot1 | plot2) + plot_annotation(tag_levels = 'A')
patch2 <- (plot3 | plot4) + plot_annotation(tag_levels = 'A')

# Save it  as a PDF
ggsave(
  filename = "../data/plot_patched_0.pdf",
  plot = patch0,
  width = 10,
  height = 12,
  units = "in"
)
ggsave(
  filename = "../data/plot_patched_1.pdf",
  plot = patch1,
  width = 16,
  height = 8,
  units = "in"
)
ggsave(
  filename = "../data/plot_patched_2.pdf",
  plot = patch2,
  width = 12,
  height = 8,
  units = "in"
)

# Figure legends ----
title1
title2
title3
title4

# Metrics ----
length(papers$ids)
papers$ids
papers[["QueryTranslation"]]

# Pubmed URLs ----
# Concatenate the main URL with each PMID
pmid_urls <-
  paste0("https://pubmed.ncbi.nlm.nih.gov/", papers$ids)

# Print all the URLs
print(pmid_urls)

# Print all the URLs without list numbers and quotes
cat(pmid_urls, sep = "\n")

# Or get pubmed IDs to paste to Zotero
cat(papers$ids,  sep = ", ")
