## Supplementary figures and images for "Reply to Dages, et al. You AIn’t using it right: Artificial intelligence progress in allergy"

### abstracts_heatmap.pdf

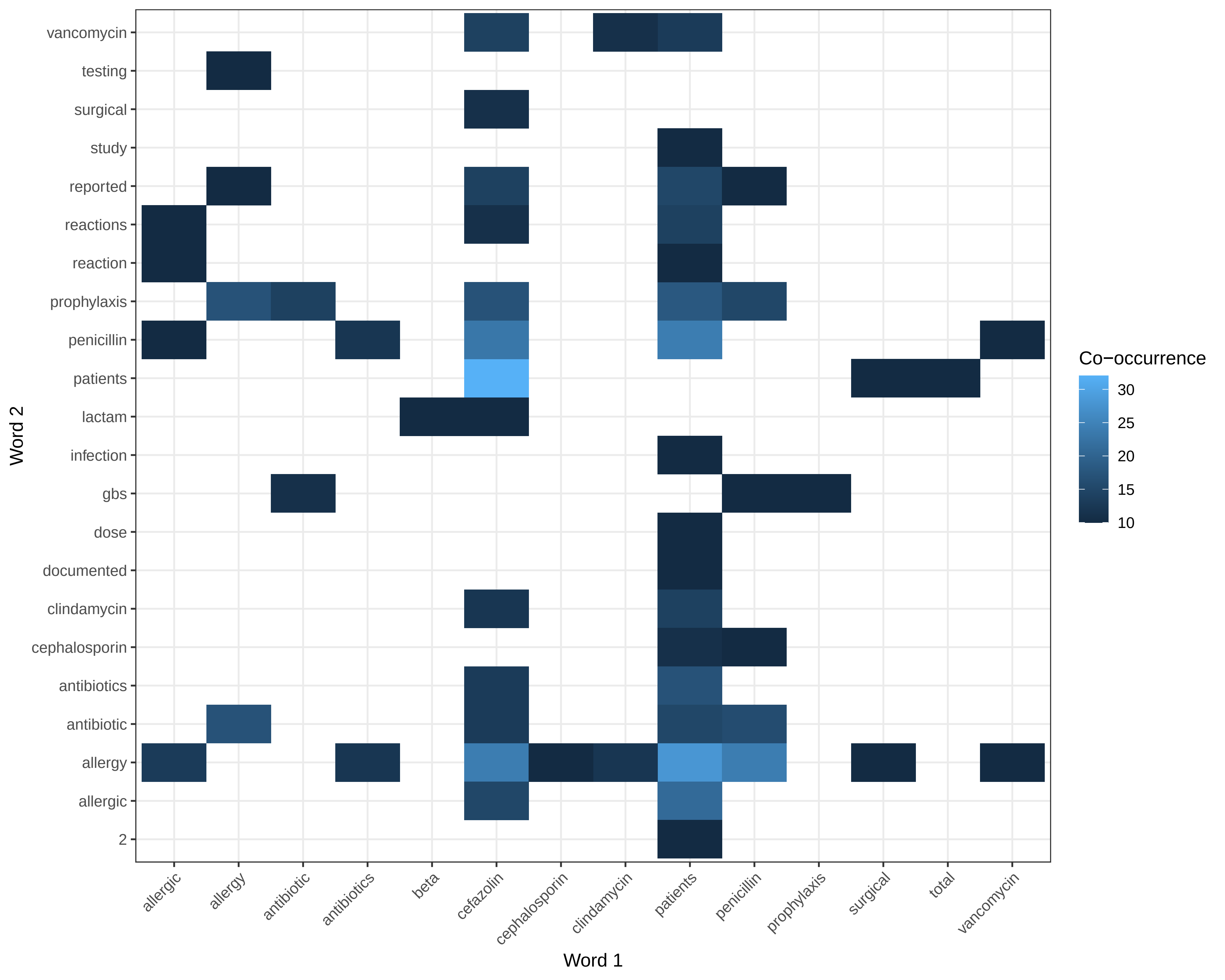

### abstracts_tf_idf.pdf

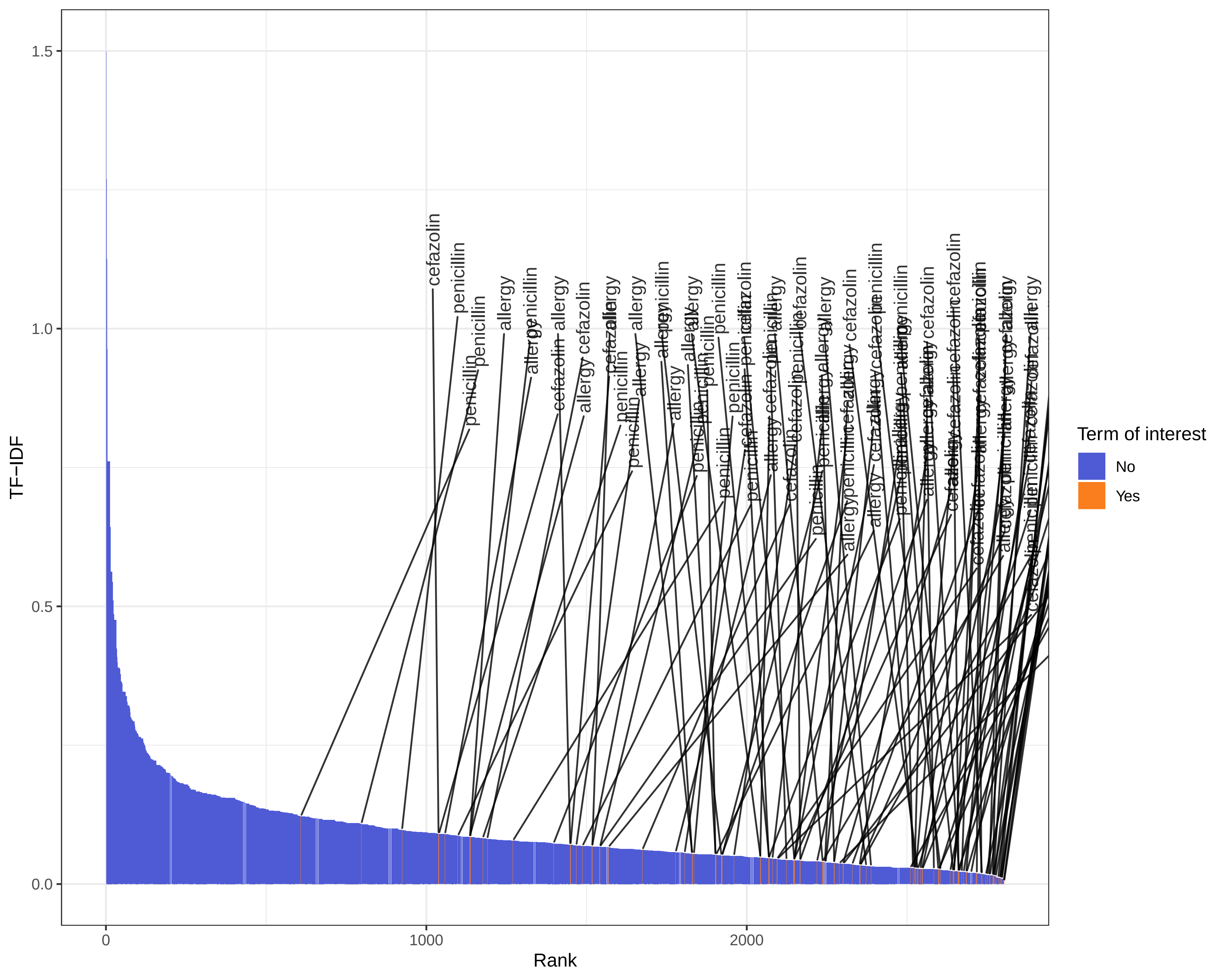

### abstracts_word_frequency.pdf

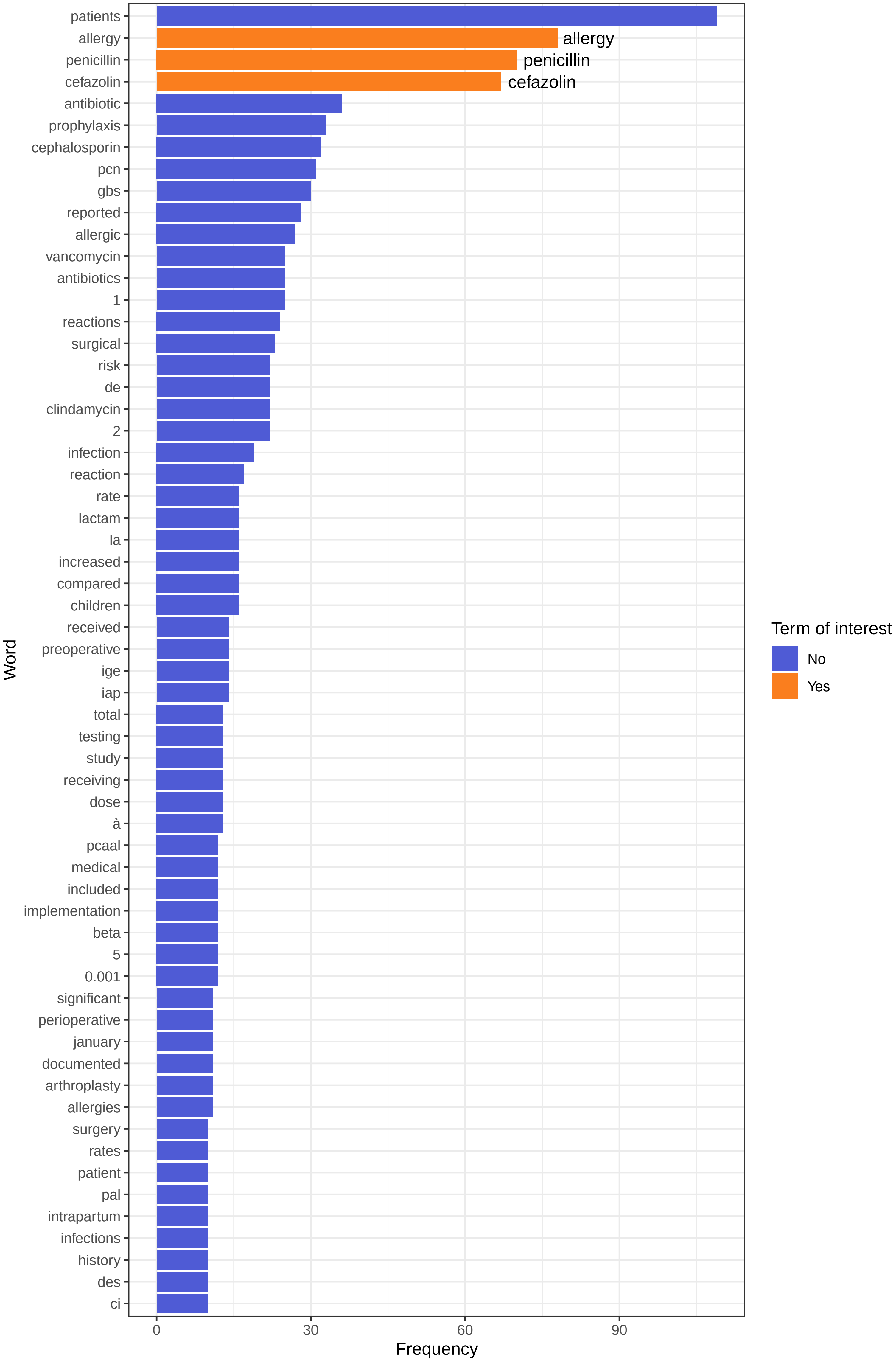

### figure_1.png

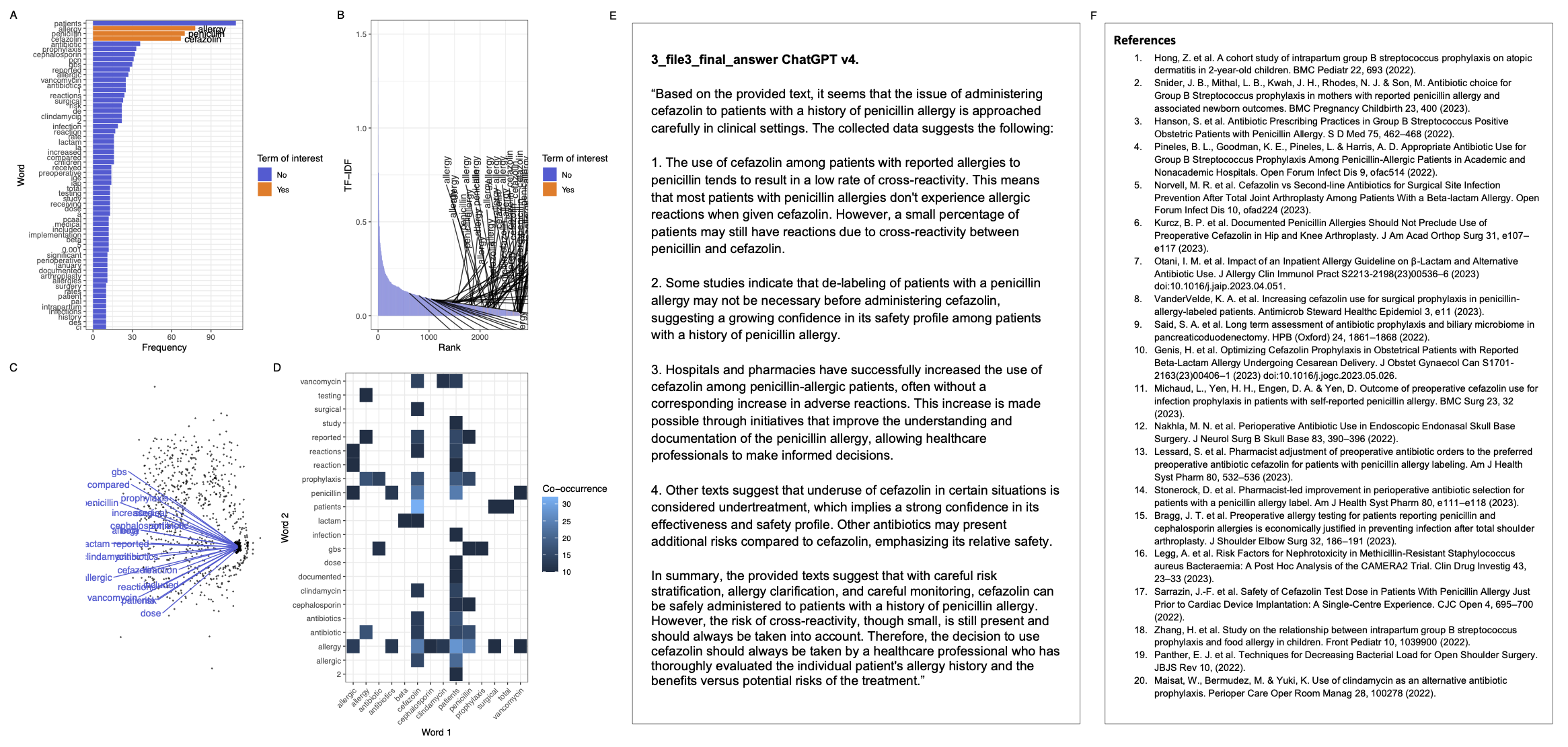

### figure_1.pptx

#### Slide 1
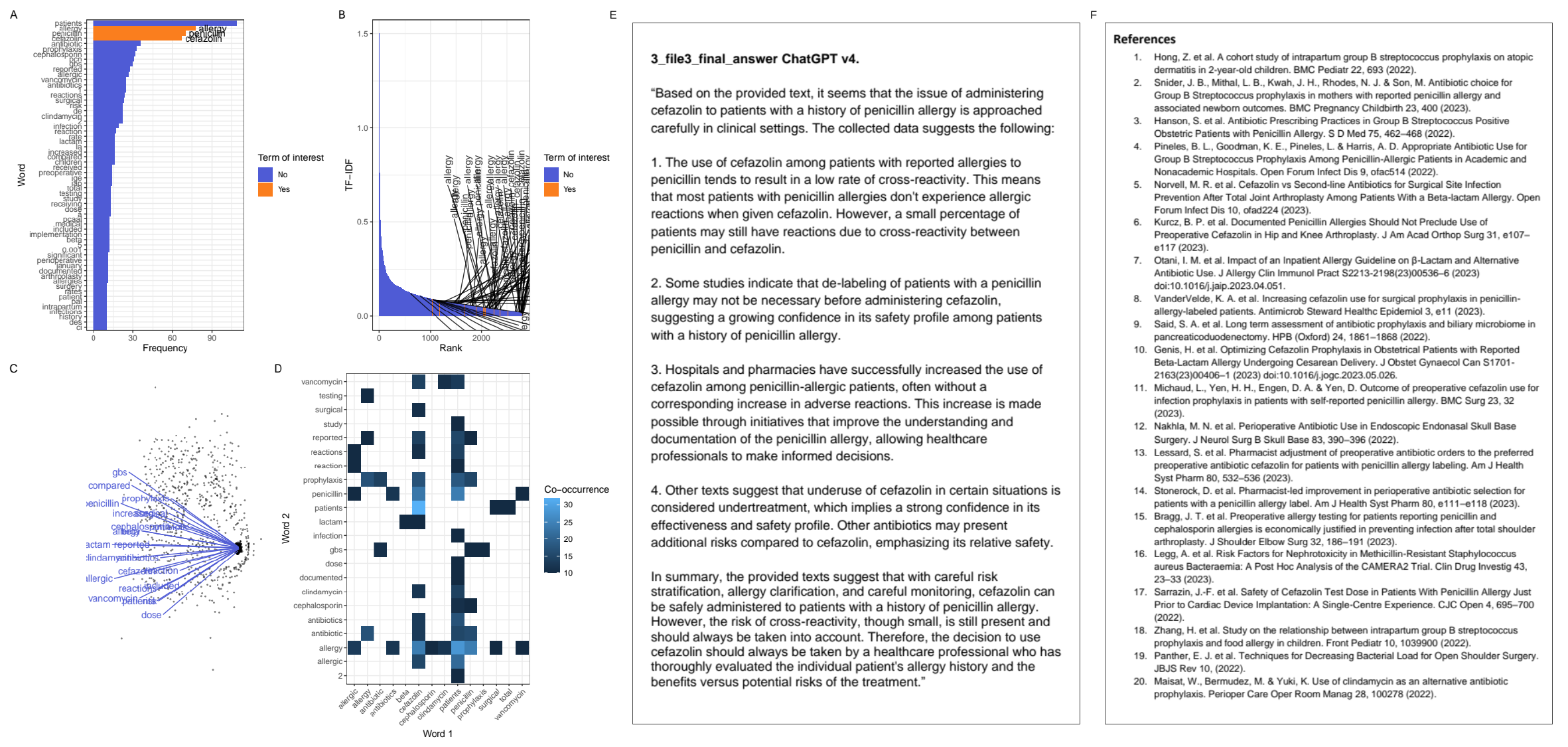

E
F

### network_plot.pdf

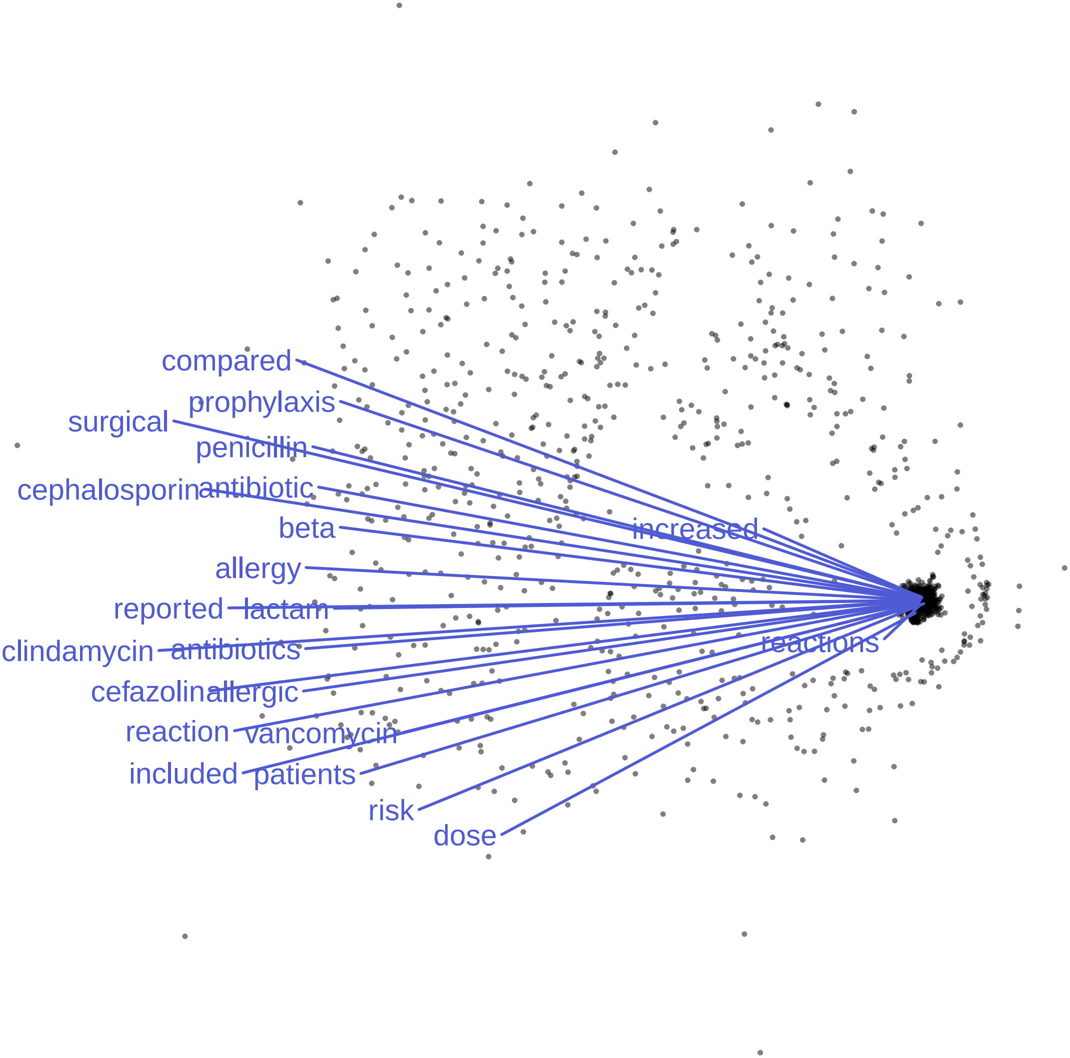

### plot_patched_0.pdf

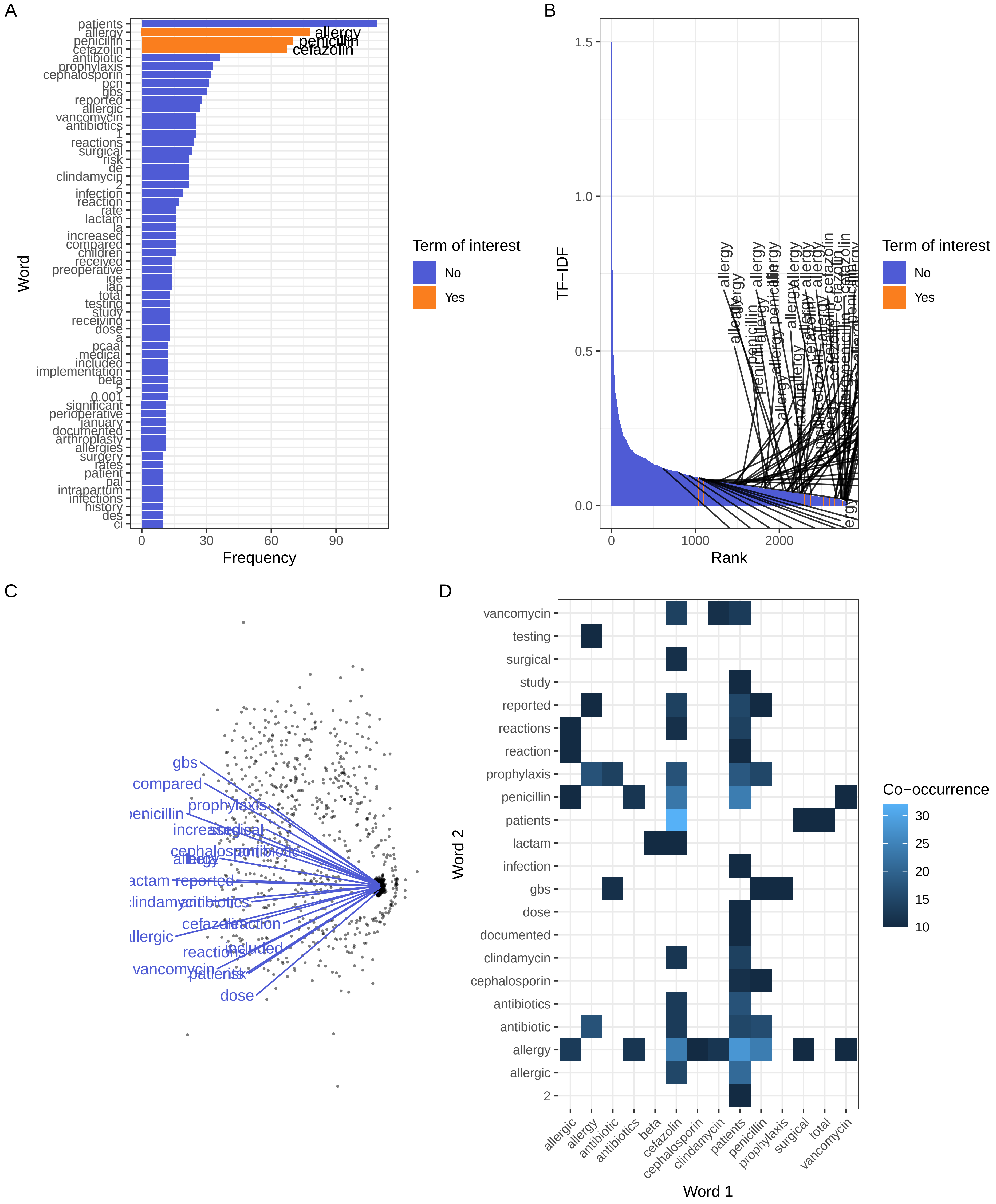

### plot_patched_1.pdf

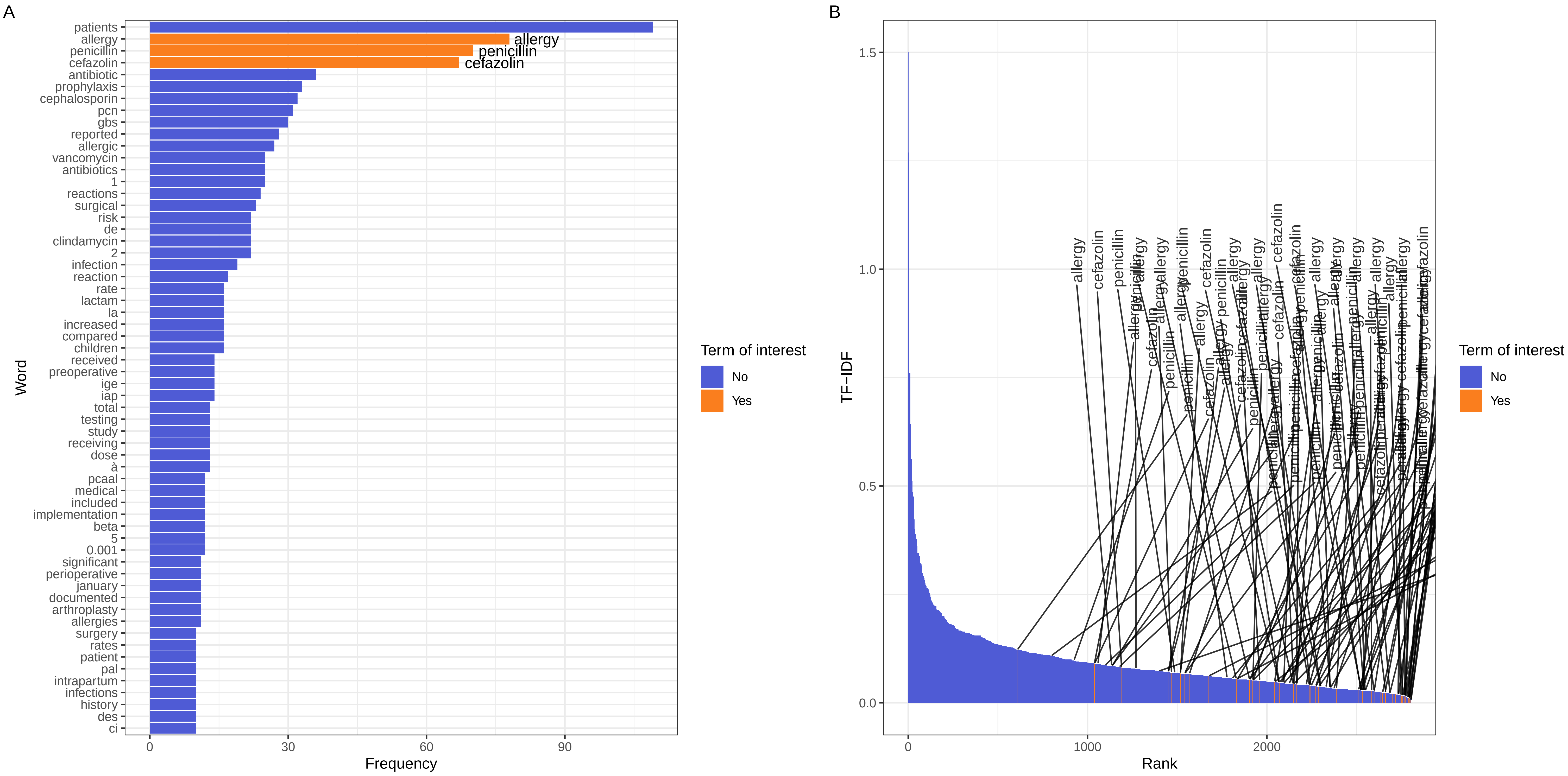

### plot_patched_1.png

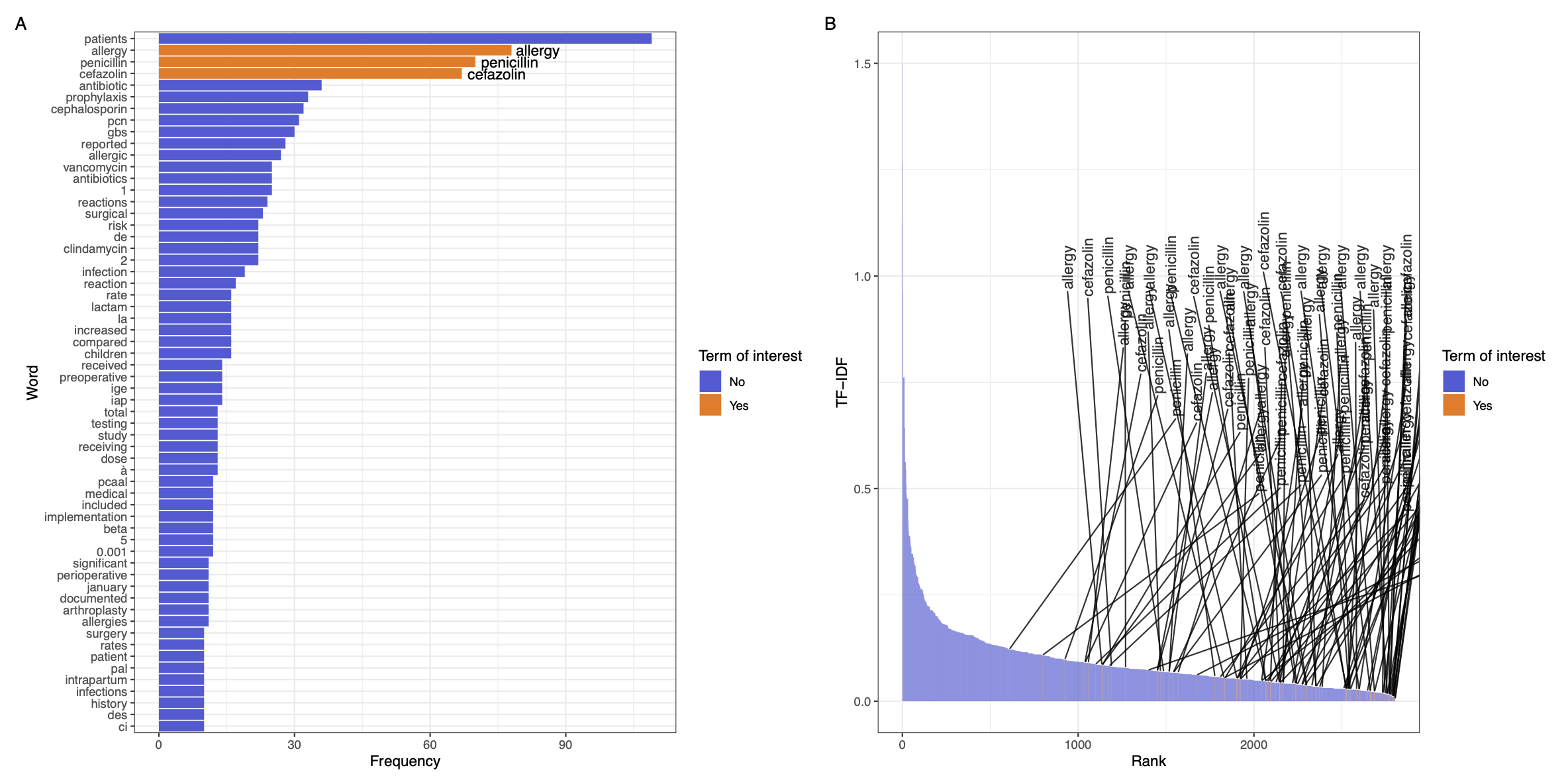

### plot_patched_2.pdf

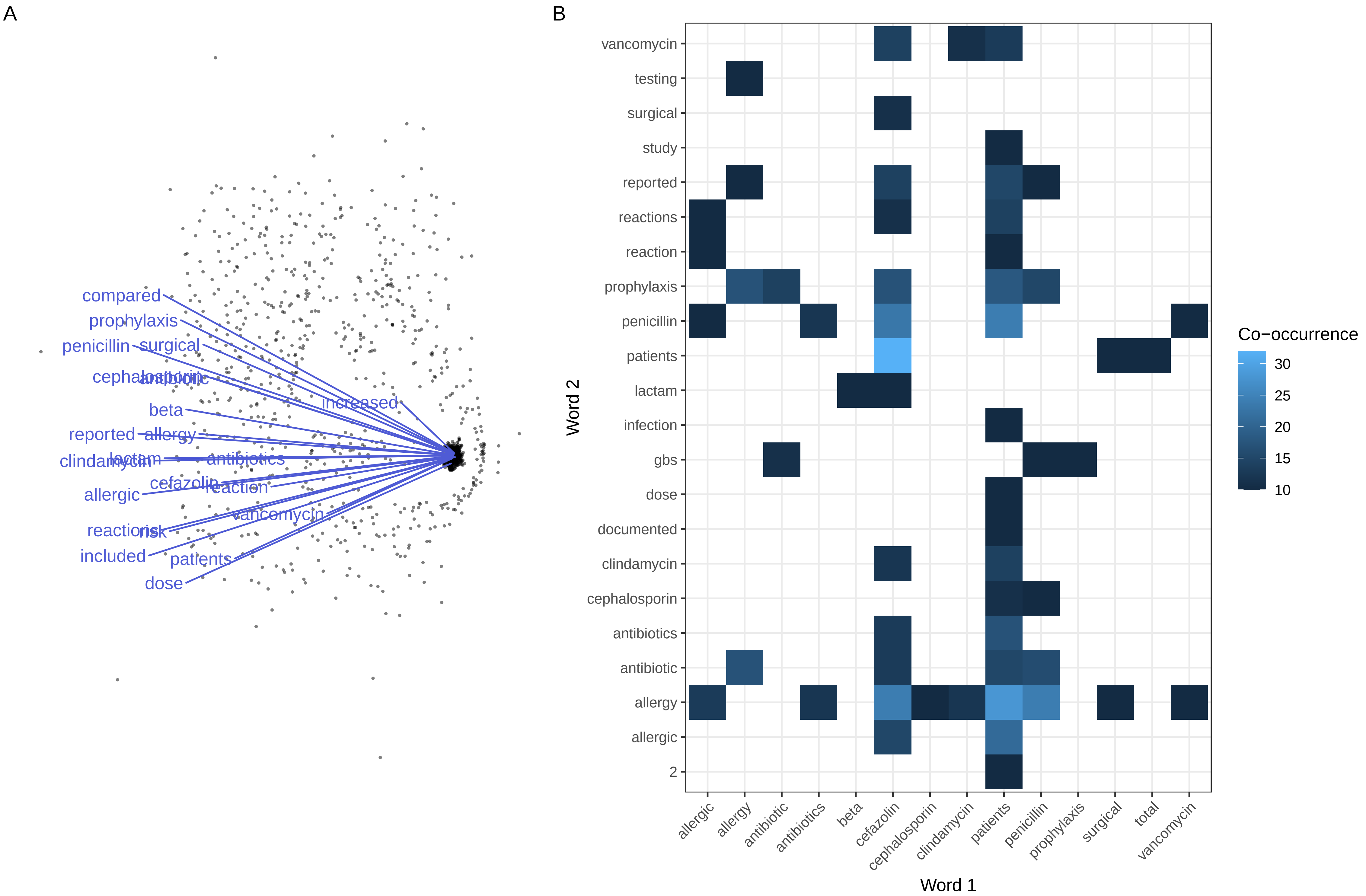

### plot_patched_2.png

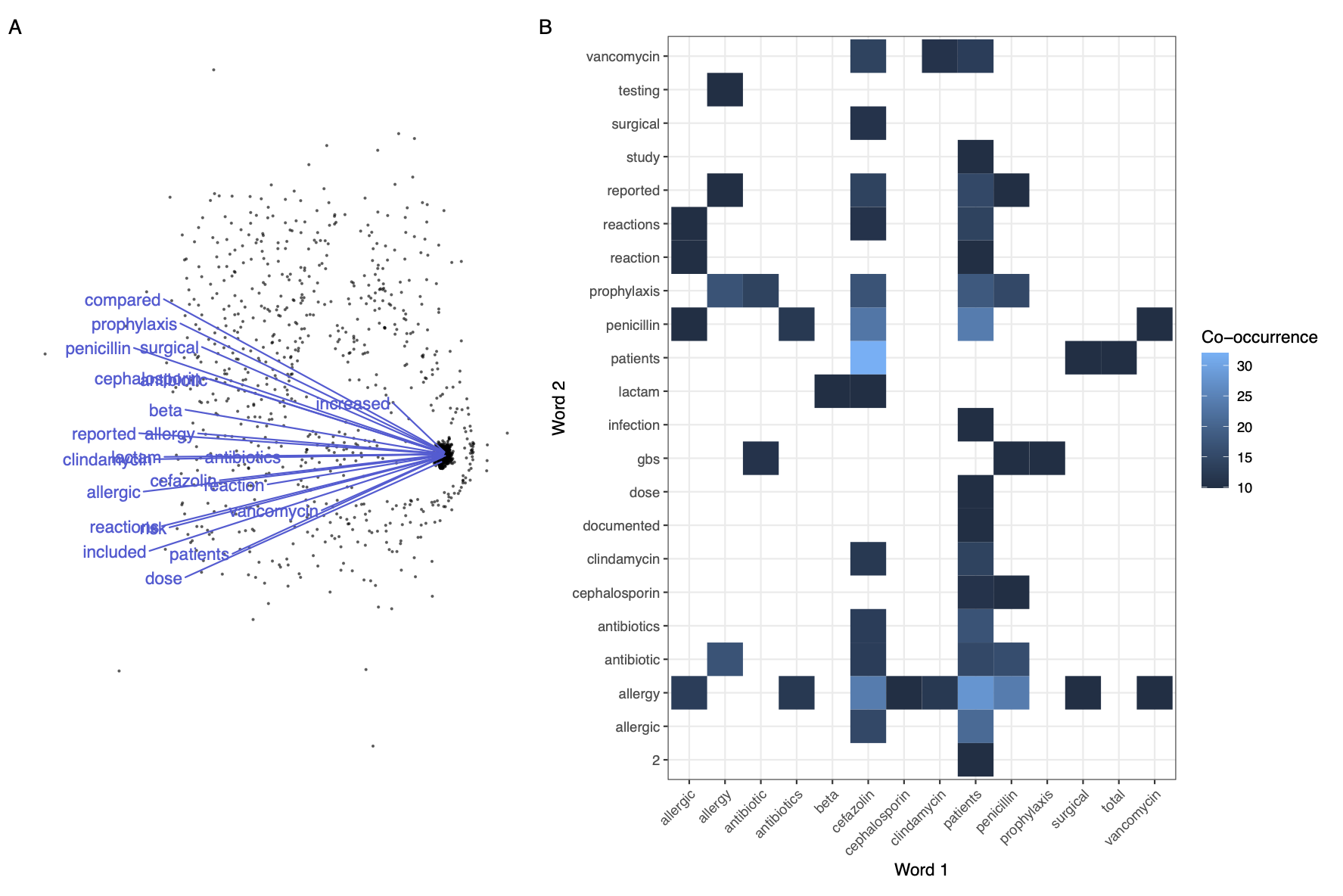
